# The clinical utility of functional testing in fibroblasts to diagnose primary mitochondrial disease

**DOI:** 10.64898/2026.06.12.26355546

**Authors:** Johan L.K. Van Hove, Marisa W. Friederich, Roxanne A. Van Hove, Jessica C. Lee, Kaz M. Knight, Tonia E. Donovan, Lori Silveira, Rebecca D. Ganetzky, Michio Hirano, Jose Abdenur, Merlin G. Butler, David Cassiman, Bruce H. Cohen, Sarah H. Elsea, Gregory M. Enns, William A. Gahl, Ralitza Gavrilova, Gabrielle C. Geddes, Emma E. Glamuzina, Amy Goldstein, Richard H. Haas, Aneal Khan, Kimberly A. Kripps, Austin A. Larson, April N. Lehman, Uta Lichter-Konecki, Johannes A. Mayr, Eva Morava, James T. Peterson, Jill A. Rosenfeld, Russell P. Saneto, Fernando Scaglia, Emily Shelkowitz, Mariella Simon, Joél E.G. Smet, Wendy E. Smith, Claudia Soler-Alfonso, Mark Tarnopolsky, Rudy N.A. Van Coster, Arnaud V. Vanlander, Pieter Vermeersch, Jerry Vockley, Kristen Wigby, Lynne A. Wolfe, Saskia B. Wortmann, Jennifer H. Yang

## Abstract

Genome sequencing of the heterogeneous primary mitochondrial disorders (PMD) frequently reveals variants of uncertain significance that require functional tests for diagnosis, and does not identify variants in all patients. We analyzed mitochondrial enzyme assays, blue native polyacrylamide gel electrophoresis (BN-PAGE) with in-gel activity staining, complex I assembly blot, and select protein abundances in fibroblasts of a case series of 204 PMD patients divided into functional classes, in comparison to 51 controls and 53 differential diagnostic conditions. Overall, sensitivity and specificity for respiratory chain enzyme assays were 46% and 93% respectively, for BN-PAGE 40% and 98%, for complex I assembly assay 49% and 99%. The overall sensitivity of all tests was 76%, specificity 93%, with positive predictive value 96% and negative predictive value 67%. Categories with high sensitivity were isolated complex deficiencies, nuclear DNA-encoded mitochondrial protein synthesis defects, co-factor defects, and mitochondrial amino-acyl-tRNA synthetase conditions when aided by protein abundance. Mitochondrial DNA mutations and maintenance disorders showed poor sensitivities. Secondary dysfunctions were rare. A complete battery of functional tests showed strong diagnostic clinical utility in fibroblasts.

**One sentence summary:** A combination of four mitochondrial functional tests to identify or confirm suspected primary mitochondrial disease in fibroblasts had good sensitivity and excellent specificity, well beyond what was perceived using enzyme assays only.

## Introduction

Mitochondria are highly dynamic organelles of tubular structure that perform many critical functions within the cell including, but not limited to, regulation of cellular metabolism, Ca^2+^ and Fe^2+^ homeostasis, steroid synthesis, lipoic acid synthesis, iron-sulfur cluster biosynthesis, detoxification of ammonia, cell cycle regulation, and apoptosis or programmed cell death. The most prominent function of mitochondria is the generation of ATP through the process of oxidative phosphorylation (OxPhos). The machinery involved in the generation of cellular energy in mitochondria is extremely efficient but highly complex. Biochemical pathways derive energy from metabolic substrates such as glucose and fat by generating reduced equivalents (e.g. NADH), which in the OxPhos pathway generate ATP while consuming oxygen. The respiratory chain that makes up the OxPhos pathway consists of five complexes composed of 91 proteins, of which 13 are encoded by the mitochondrial genome. All the remaining proteins are encoded by the nuclear genome and imported into the mitochondria (Frazier et al, 2019; Rahman et al. 2018; Hock et al 2020). The biogenesis of the five complexes into higher order structures follows an intricate assembly process that depends on precise and coordinated cytosolic and mitochondrial translation, resulting in the appropriate stoichiometric levels of its subunits (Hock et al. 2020; Barros et al. 2020; Fernández-Vizarra et al. 2009). If a single subunit protein is absent or reduced, then the entire complex may not assemble properly, creating a co-dependency within mitochondrial complexes as elegantly illustrated for complex I (Hock et al., 2020; Stroud DA et al. 2016). The 13 mitochondrial DNA-encoded subunits of these complexes are transcribed and translated on the mitochondrial ribosomal machinery prior to their assembly into the complexes. Disorders can arise at each step of the mitochondrial biogenesis, resulting from pathogenic variants in mitochondrial DNA (mtDNA) or in hundreds of nuclear DNA (nDNA) encoded genes (Frazier et al.,2019; Rahman et al., 2018; Hock DH et al., 2020; Thompson et al. 2020; Gusic et al., 2021; Stenton et al, 2018; Stenton et al., 2020; Schlieben & Prokisch, 2020).

Genetic disorders of the OxPhos system result in primary mitochondrial disease (PMD) (Frazier et al.,2019; Rahman et al., 2018; Hock DH et al., 2020; Thompson et al. 2020; Gusic et al., 2021; Stenton et al, 2018; Stenton et al., 2020; Schlieben & Prokisch, 2020; Koopman et al. 2012; Gorman et al. 2016; Rius et al. 2025) Primary mitochondrial disease is the most common cause of metabolic disease with a frequency of at least 1:5,000 (Gorman et al., 2015; Schaefer et al., 2008; Skladal et al., 2003). They are characterized by extensive biochemical, clinical, and genetic heterogeneity with pathogenic variants in over 360 genes identified(Frazier et al.,2019; Rahman et al., 2018; Hock DH et al., 2020; Thompson et al. 2020; Gusic et al., 2021; Stenton et al, 2018; Stenton et al., 2020; Schlieben & Prokisch, 2020; Koopman et al. 2012; Gorman et al. 2016; Rius et al. 2025). The resulting extensive phenotypic heterogeneity makes it difficult to recognize patients that require diagnostic testing, which is often challenging, lengthy, and frustrating, with long delays serving as a barrier to disease-specific care (Grier et al., 2018; Thompson et al., 2023A). A molecular diagnosis is considered essential (Parikh et al, 2019). Upon clinical and radiological indications, sometimes aided by biomarkers, the current first line diagnostic investigation is next generation sequencing (next-gen) of mitochondrial DNA and of the exome or genome, which provides a putative diagnostic result in 50 to 65% of mitochondrial patients (Figure 1) (Thompson et al., 2020; Gusic et al., 2021; Stenton et al., 2018; /Stenton et al., 2020; Schlieben et al., 2020; Koopman et al., 2012; Gorman et al., 2016; Rius et al., 2025Wortmann et al., 2015; Taylor et al, 2014; Lieber et al, 2013). While effective, several problems persist, including frequent variants of uncertain significance (VUS), which require validation by functional assays for definitive diagnostic interpretation. The 35% of patients with non-revealing next-gen sequencing studies require functional studies to guide subsequent advanced molecular studies. Past diagnostic practice involved measurement of respiratory chain enzyme (RCE) activities in muscle or liver biopsies (Munnich et al., 1996; Rustin et al. 1994; Thorburn et al., 2001; Thompson et al, 2023B). However, these assays do not provide a complete analysis of mitochondrial function, are invasive, and secondary dysfunction due to nongenetic factors causes problems of specificity. Skin biopsy for fibroblast culture is a minimally invasive, in-office procedure, preferred by patients and physicians. Retrospective analysis of RCE activities in fibroblasts had shown a sensitivity of only ∼50% of cases (Thorburn et al, 2001; Thompson et al., 2023B; Niers et al, 2003; Faivre et al., 2000; Robinson et al., 1990). but systematic studies in the molecular era have not been available. Further, new functional assays applicable for use in fibroblasts have been developed to improve the evaluation of mitochondrial function.

**Figure 1:**
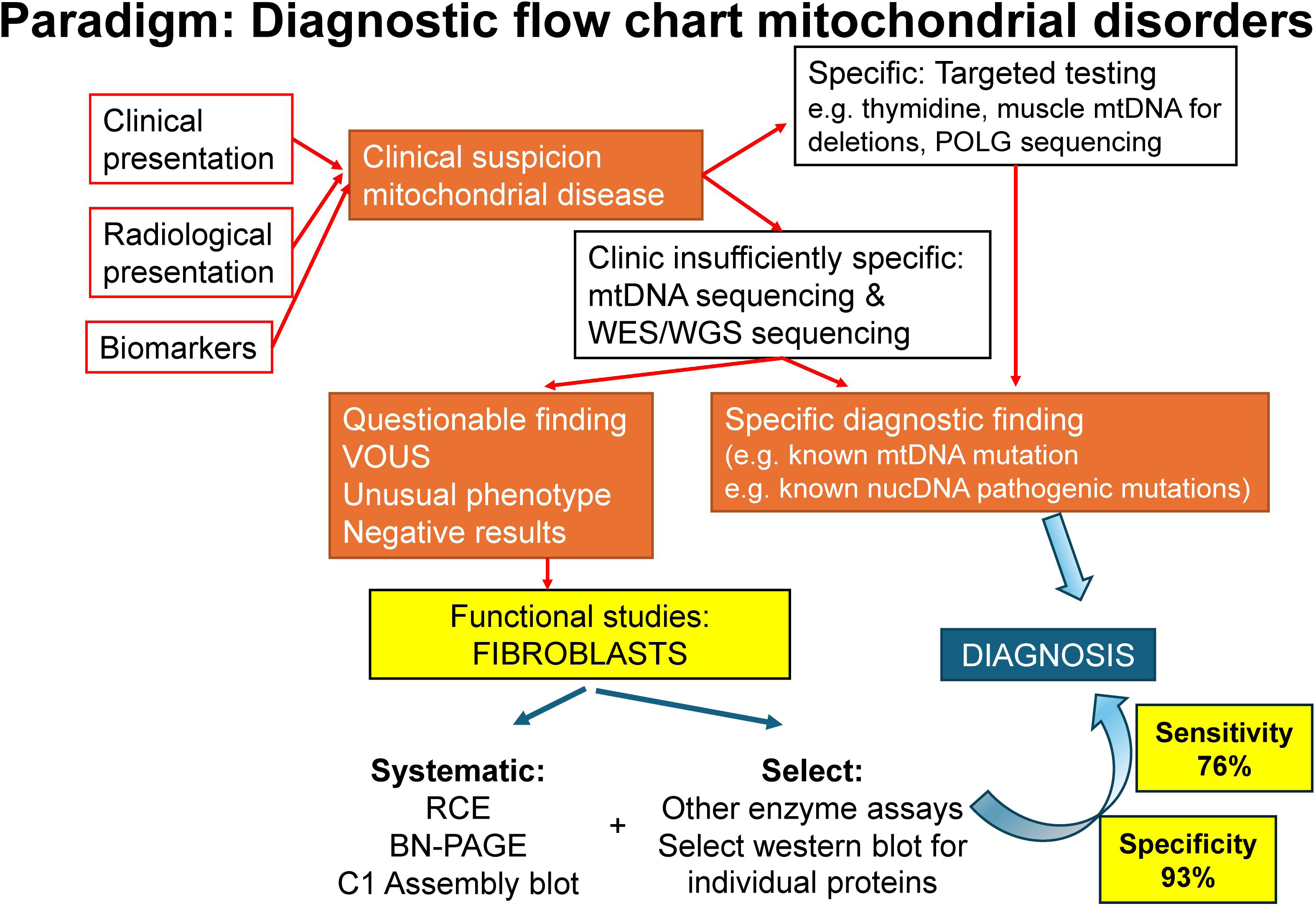
Paradigm for the diagnosis of Primary mitochondrial disorders. Upon clinical suspicion of a mitochondrial disorder, aided by radiological and biomarker studies, molecular testing constitutes first line testing. When next-gen sequencing returns variants of uncertain specificity (VUS) or does not identify a candidate result, then mitochondrial functional testing are needed. In this study, we routinely employed respiratory chain enzyme assay, blue native polyacrylamide gel electrophoresis (BN-PAGE) with in-gel activity staining and complex I assembly assay, sometimes aided by specific abundances of pertinent proteins. Our study showed that diagnostic findings were returned in fibroblasts with a sensitivity of 76% and a specificity of 93%.

Therefore, we applied a series of functional testing methods (enzyme assays, blue native polyacrylamide gel electrophoresis (BN-PAGE) with in-gel activity staining, and complex I assembly assay, and as indicated protein abundance by western blot) to a case series of primary mitochondrial disease patients and controls and established their clinical utility for the diagnosis and confirmation of primary mitochondrial disease in fibroblasts derived from a skin biopsy. Given the large genetic heterogeneity inherent in this study for the analysis of functional assays the data were grouped according to the following functional categories: (1) isolated complex deficiencies due to defect in subunits or assembly factors (complex I, complex II, complex III, complex IV, complex V, or pyruvate dehydrogenase (PDH)), (2) co-factor deficiencies (coenzyme Q, lipoate, iron-sulfur cluster, S-adenosylmethionine (SAM)), and the combined deficiencies were subcategorized as follows (3) mitochondrial DNA pathogenic variants of tRNAs and singe large-scale mitochondrial DNA deletions, (4) mitochondrial DNA maintenance disorders, (5) the mitochondrial amino-acyl tRNA synthetases (ARS2 disorders including the Gat^CAB^ complex) (Friederich et al., 2018), (6) mitochondrial protein synthesis disorders including disorders of mitochondrial RNA transcription, processing, translation on the mitoribosome and mitochondrial protein importation, and (7) mitochondrial structure (fusion and fission) disorders. For comparison, we included normal control fibroblasts, but for specificity, also included differential diagnostic conditions, including (1) metabolic disorders with emphasis on those with potential secondary mitochondrial dysfunction, (2) neuromuscular disorders with potential for clinically overlapping symptoms, and (3) genetic disorders emphasizing disorders affecting gene regulation. We showed that testing in fibroblasts can be optimized using an appropriate combination of tests with clinical robustness, high sensitivity and specificity to allow for effective mitochondrial functional evaluation in patients with primary mitochondrial disease.

## Results

### Cell lines

For this study, we analyzed 308 fibroblast cell lines (Table EV1). There were 204 cell lines from subjects with the following distribution of primary mitochondrial disorders: 88 isolated complex deficiencies (39 complex I, six complex II, four complex III, 12 complex IV, 16 complex V and 11 PDH), 102 combined deficiencies (20 mtDNA variants, 15 mtDNA maintenance disorders, 40 ARS2 defects, and 28 mitochondrial protein synthesis disorders comprised of seven RNA processing defects, 20 translation defects and 1 protein import disorder), 11 co-factor defects (one coenzyme Q, four lipoate, five iron-sulfur clusteropathies, and one SAM import Kishita et al., 2015)), and two mitochondrial structural disorders. There were 53 differential diagnostic cell lines from subjects with the following disorders: 32 metabolic diseases, 11 myopathies or neuropathies, and 10 genetic syndromes, and 51 normal control cell lines. For all cell lines, basic assays of RCE, BN-PAGE with in-gel activity staining, and complex I assembly blot were completed (Table EV2 and Table EV3).

### Cutoff optimization

The cutoff was evaluated by minimizing false positives in normal controls while maximizing sensitivity. The lowest value observed in any controls was at 46% to 50% of average controls, and at −2.48 to −0.23 SD for the Z-score (Table EV4). Similarly, for the ratios of the complex enzyme activity over citrate synthase activity, all values remained above 47% of average controls and −2.12 SD Z-score. When we evaluated the sensitivity and specificity across cutoffs, an increase was observed in sensitivity between cutoff 44% and 45% of average controls for complex I (from 0.23 to 0.25), an increase in sensitivity of complex III between 41% and 42% (from 0.4 to 0.6), a gradual increase for complex IV between 41% and 49% (from 0.21 to 0.29), with no impact on complex II in this range (Appendix Table 2). For use in a clinical laboratory, we set the optimized cutoff at 45% of average of controls which maximized sensitivity for complexes I and IV while maintaining all control samples in the normal range. Similarly, for the Z-score, complex I had an increase in sensitivity between Z-score cutoff −2.7 to −2.5 SD from 0.22 to 0.25, complex II had an equal increase between Z-score −2.7 and −2.5 SD from 0.58 to 0.67, with complex III showing an increase in sensitivity between −2.9 and −2.7 SD, and for complex IV we again noted a gradual increase at all levels between −2.7 and −2.1 SD (Appendix Table3). We set the optimized cutoff of the Z-score at −2.5 SD, which optimized sensitivity for all complexes (except IV) while maintaining all control samples in the normal range. A positive result was defined as either <40% of controls or, for a “borderline result”, a combination of <45% and <-2.5 SD for the activity as well as for the ratio of the activity over citrate synthase. For the complex V hydrolytic assay, the observed normal range extended to Z-score −1.8 SD or 68% of median of controls. For robustness, we identified that there was no impact of passage number up to 15 passages longitudinally (ANOVA p>0.1 for each assay), and no impact of the presence or not of antibiotics and antimycotics (paired Student t-test p>0.2 on each enzyme assay and ratio).

### Diagnostic utility

We next reviewed diagnostic utility for each specific diagnostic category.

### Isolated complex deficiencies (Table 1)

For isolated complex I deficiency, there were nine cell lines with pathogenic variants in mtDNA-encoded subunits, and 30 with variants in nuclear DNA-encoded subunits. The RCE had a sensitivity of 54% (6/9 for mtDNA encoded and 15/30 for nuclear DNA encoded). The BN-PAGE had poor sensitivity of 36% (0% in mtDNA-encoded subunits), but the complex I assembly blot had strong sensitivity 79%, particularly for nuclear DNA-encoded disease (normal in 5/9 mtDNA-encoded, but normal in only 3/30 nuclear DNA-encoded cell lines). Collectively, an abnormality in any of three tests was detected in 92% of cases. In mtDNA-encoded cases, the RCE was the most effective test, whereas in nuclear DNA-encoded cases, the complex I assembly blot was the most effective test. The only nuclear DNA-encoded case with normal results had pathogenic variants in *NDUFAF5*, while for mtDNA-encoded cases normal results were limited to pathogenic variants in *MT-ND6* associated with Leber hereditary optic neuropathy.

**Table 1:**
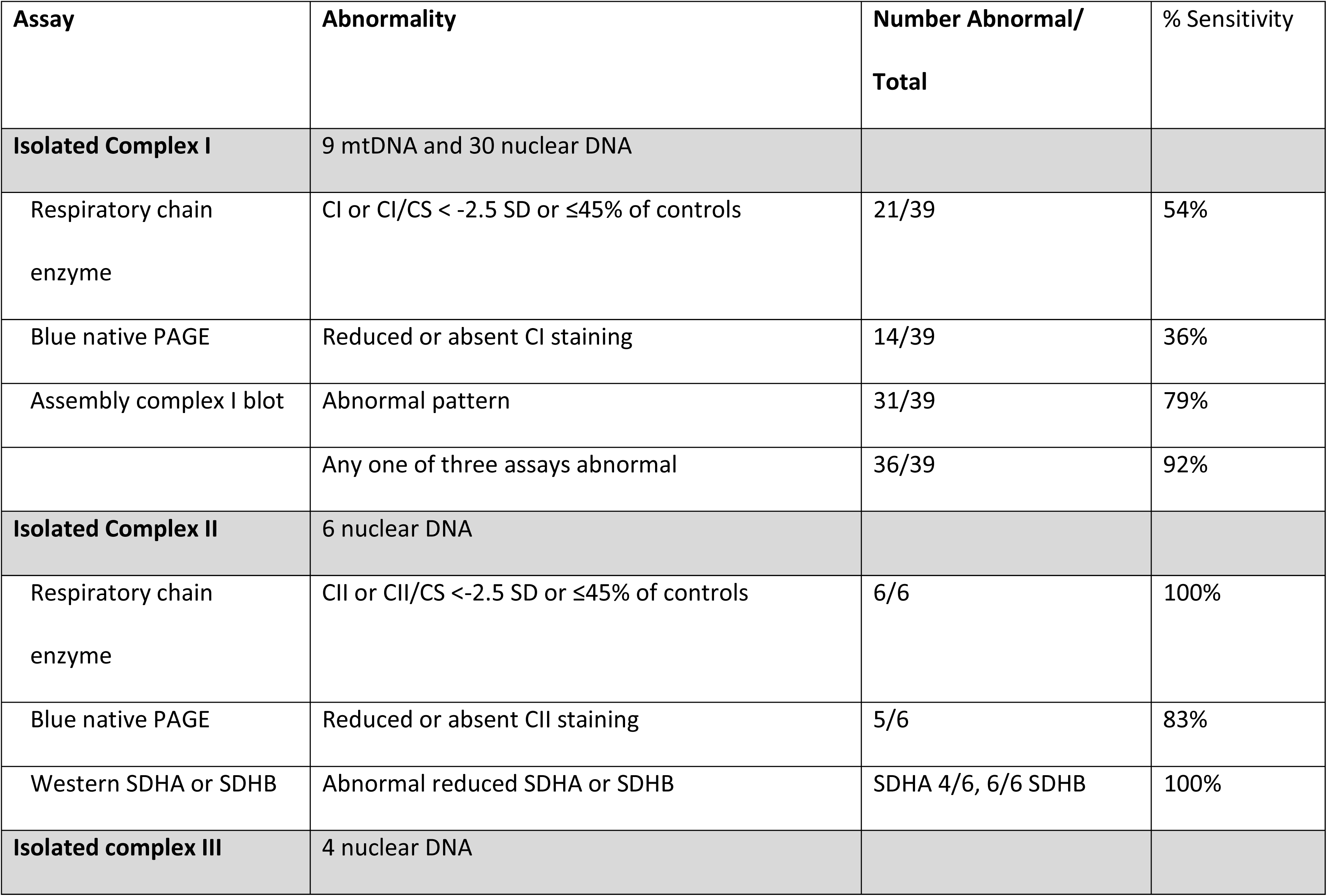
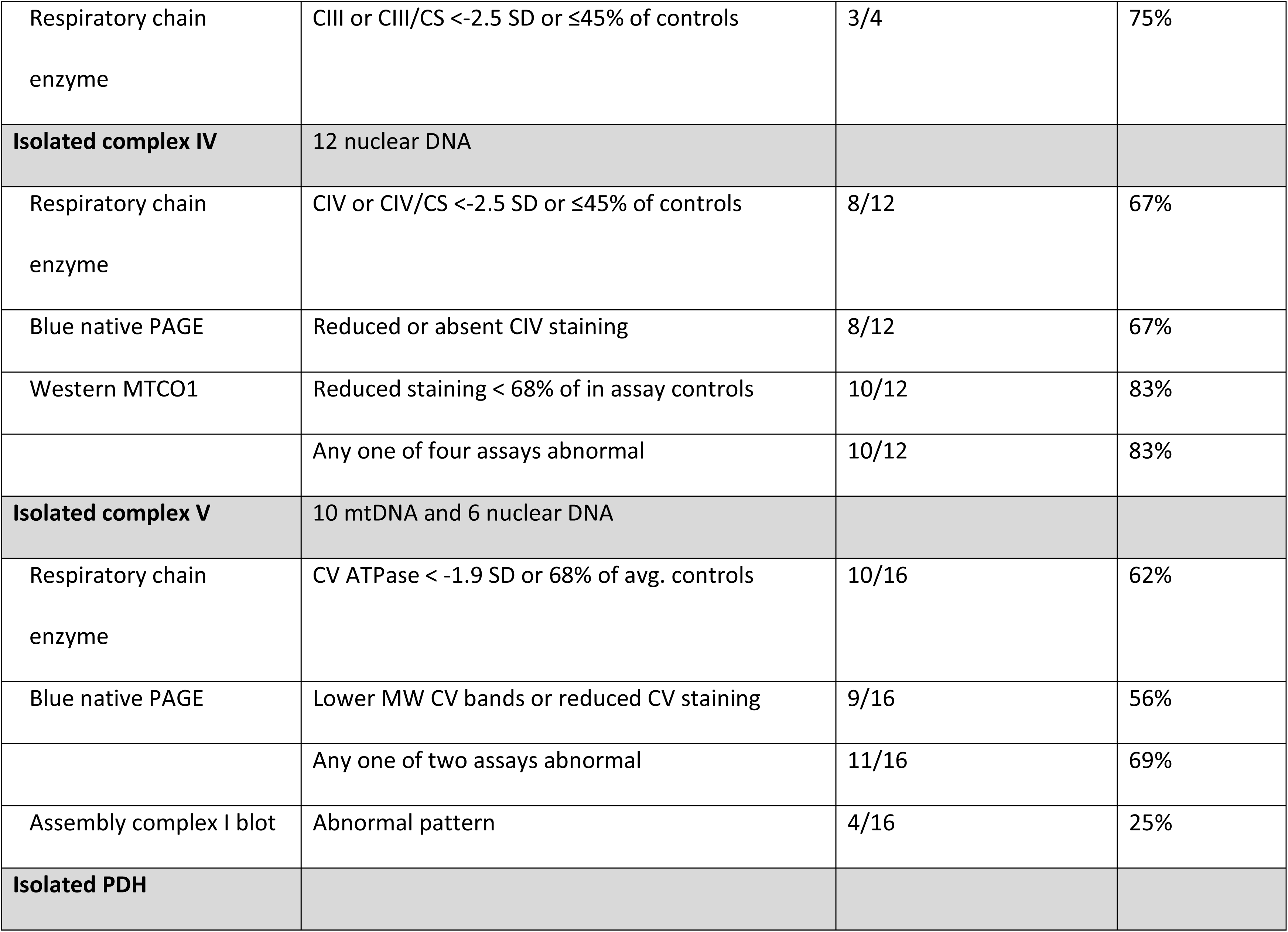
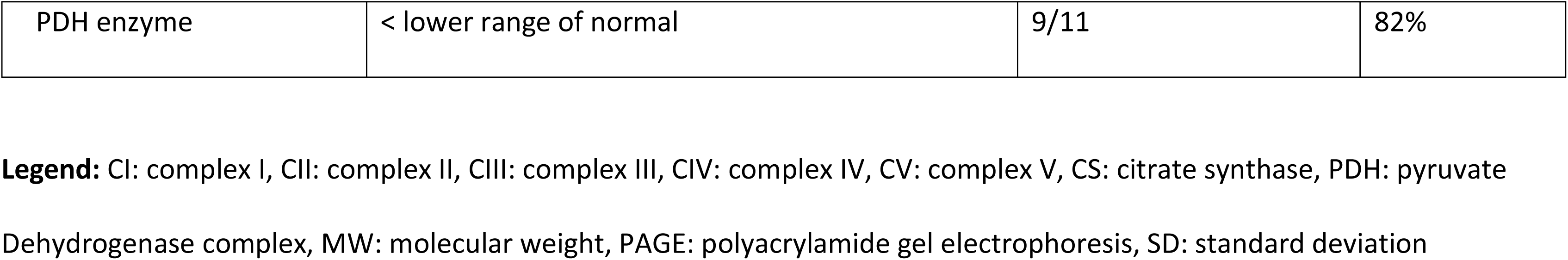
Isolated complex deficiencies.

| Assay | Abnormality | Number Abnormal/<br>Total | % Sensitivity |
| --- | --- | --- | --- |
| **Isolated Complex I** | 9 mtDNA and 30 nuclear DNA |  |  |
| Respiratory chain<br>enzyme | CI or CI/CS < -2.5 SD or ≤45% of controls | 21/39 | 54% |
| Blue native PAGE | Reduced or absent CI staining | 14/39 | 36% |
| Assembly complex I blot | Abnormal pattern | 31/39 | 79% |
|  | Any one of three assays abnormal | 36/39 | 92% |
| **Isolated Complex II** | 6 nuclear DNA |  |  |
| Respiratory chain<br>enzyme | CII or CII/CS < -2.5 SD or ≤45% of controls | 6/6 | 100% |
| Blue native PAGE | Reduced or absent CII staining | 5/6 | 83% |
| Western SDHA or SDHB | Abnormal reduced SDHA or SDHB | SDHA 4/6, 6/6 SDHB | 100% |
| **Isolated complex III** | 4 nuclear DNA |  |  |
| Respiratory chain enzyme | CIII or CIII/CS <-2.5 SD or ≤45% of controls | 3/4 | 75% |
| **Isolated complex IV** | 12 nuclear DNA |  |  |
| Respiratory chain enzyme | CIV or CIV/CS <-2.5 SD or ≤45% of controls | 8/12 | 67% |
| Blue native PAGE | Reduced or absent CIV staining | 8/12 | 67% |
| Western MTCO1 | Reduced staining < 68% of in assay controls | 10/12 | 83% |
|  | Any one of four assays abnormal | 10/12 | 83% |
| **Isolated complex V** | 10 mtDNA and 6 nuclear DNA |  |  |
| Respiratory chain enzyme | CV ATPase < -1.9 SD or 68% of avg. controls | 10/16 | 62% |
| Blue native PAGE | Lower MW CV bands or reduced CV staining | 9/16 | 56% |
|  | Any one of two assays abnormal | 11/16 | 69% |
| Assembly complex I blot | Abnormal pattern | 4/16 | 25% |
| **Isolated PDH** |  |  |  |
| PDH enzyme | < lower range of normal | 9/11 | 82% |

**Legend:** CI: complex I, CII: complex II, CIII: complex III, CIV: complex IV, CV: complex V, CS: citrate synthase, PDH: pyruvate
Dehydrogenase complex, MW: molecular weight, PAGE: polyacrylamide gel electrophoresis, SD: standard deviation

For isolated complex II deficiency, the RCE was abnormal in all cases (100% sensitivity), whereas BN-PAGE was abnormal in 5/6 cases. The abundance of SDHA was decreased in 4/6 cases, whereas the abundance of SDHB was decreased in all six cases including cases with mild SDHA variants, likely reflecting the need for intact SDHA to assembly SDHB. There were no false positive complex I assembly blots. For isolated complex III cases, the RCE was abnormal in three of four cases for the direct complex III assay (75%) (but not for the linked succinate:cytochrome coxidoreductase II-III assay), with no false positive cases on BN-PAGE. Complex I appeared normal, but a faint band at 950 kDa was sometimes visible (Blue et al., 2024), compatible with the need for complex I to associate with complex III in the growing supercomplex formation to finalize the last step of its assembly with the addition of the N-module (Letts et al., 2019; Protasoni et al., 2020; Acin-Perez et al., 2004). One case with pathogenic variants in *UQCRC2* had normal complex III RCE activity, but proteomics analysis showed abnormal complex III abundance (Blue et al., 2024).

For isolated complex IV deficiency, RCE was abnormal in 67% of cases, and in those cases, the BN-PAGE also showed low activity for complex IV. All cases with *SURF1* deficiency had an activity of <35% of mean of in-assay controls. The subunit MTCO1 (COX1) encoded by mtDNA gene *MT-CO1* is central to an early step in the assembly of complex IV and is sensitive to failure of early complex IV assembly. The distribution of MTCO1 protein levels on western blot was analyzed in 17 controls and ranged from 68 to 135% of the average of controls. The abundance of MTCO1 was decreased in all cases with low RCE, but also in two cases with missense variants in *SCO1* and *SCO2* respectively, confirming their diagnosis, thus providing a sensitivity of 83%. In two cases of the late assembly factor *COXF4* (Hock et al., 2020; Pitceathly et al. 2013) all assays were normal including, as expected, the MTCO1 abundance.

For isolated complex V deficiency, there were six nuclear DNA-encoded cases and 10 mtDNA encoded cases. RCE assays of complexes I through IV were normal, except for TMEM70, as expected since it also functions as an assembly factor of complex I (Sánchez-Caballero et al., 2020; Carroll et al., 2021). The hydrolytic complex V assay showed abnormal results in all nuclear DNA-encoded cases, and in four of ten mtDNA-encoded cases, for an overall sensitivity of 62%. The BN-PAGE assays showed an abnormal pattern in 56% of cases. In mtDNA-encoded cases, bands of lower MW were present. In nuclear DNA-encoded cases, the pattern reflected the impact on complex V assembly, with *ATP5F1A* and *ATP5F1D* revealing decreased holocomplex bands, whereas cells with *ATP5PO* and *TMEM70* pathogenic variants showed the presence of lower MW bands (Ganapathi et al., 2022; Fielder et al, 2025; Oláhová et al., 2018). An abnormality on either assay had a sensitivity of 69%, consisting of 100% in nuclear DNA-encoded cases but only 50% in mtDNA-encoded cases. Interestingly, four cases showed an abnormality of the assembly of complex I without abnormal RCE activity. Abnormalities in complex I in *MT-ATP6* cases have been previously noted (Morava et al., 2006), and there are shared assembly factors (Sánchez-Caballero et al., 2020; Carroll et al., 2021), and given the recurrent nature this was not considered a false positive result but a feature of this assay. On western blot analysis, in all nuclear DNA-encoded cases the abundance of ATP5PO protein was reduced and ATP5F1A protein was reduced in five of six cases, being normal in the *ATP5PO* case. ATP5F1A abundance was normal in all *MT-ATP6* cases, where ATP5PO abundance was reduced in only one case.

For isolated pyruvate dehydrogenase deficiency, the PDH enzyme activity was reduced in 82% of cases, with the normal activity as expected in two female *PDHA1* cases, which is consistent with previous reports (Shen et al., 2017; Merkevicius et al., 2025). There were no abnormalities on other assays and the lipoate western blot was also normal in three examined cases, providing no false positives.

### Combined enzyme deficiencies (Table 2)

#### mtDNA mutations

In the 20 mtDNA-encoded cases of tRNA pathogenic variants and mtDNA deletions, an abnormality on RCE was found in 25% (5/20) of cases, complex I in three cases and both complexes I and IV in two cases. An abnormality on BN-PAGE was present in 35% (7/20) of cases, most often as lower MW bands of complex V (N=5), and as an abnormality on complex I assembly assay in 40% of cases. An abnormality on either of three tests was present in 60% of cases. Progressive loss of heteroplasmy in fibroblast culture has been reported for mtDNA deletions (Bourgeron et al., 1993), therefore heteroplasmy was analyzed. For mtDNA encoded tRNA variants, of the 14 cell lines in which heteroplasmy was determined in the fibroblasts at the end of the studies, two had lost the presence of the pathogenic variant and in three cell lines was the heteroplasmy <50%, whereas in nine cell lines was the heteroplasmy in the fibroblasts >60%. Of these nine high heteroplasmy level cell lines, three samples had normal results (two with m.3243A>G at 84.3% and 85.3%, and homoplasmic *MT-TE* m.14674T>C variant). Abnormal results were present in the three of five cell lines with heteroplasmy <50%. This difference by heteroplasmy levels was not statistically significant (Fisher exact test). Of note, all but one of the *MT-ATP6* variant cell lines had near homoplasmy; and so heteroplasmy did not influence the results. The results of complex I studies in cells with *ND* pathogenic variants also did not appear to be related to heteroplasmy rates.

**Table 2:**
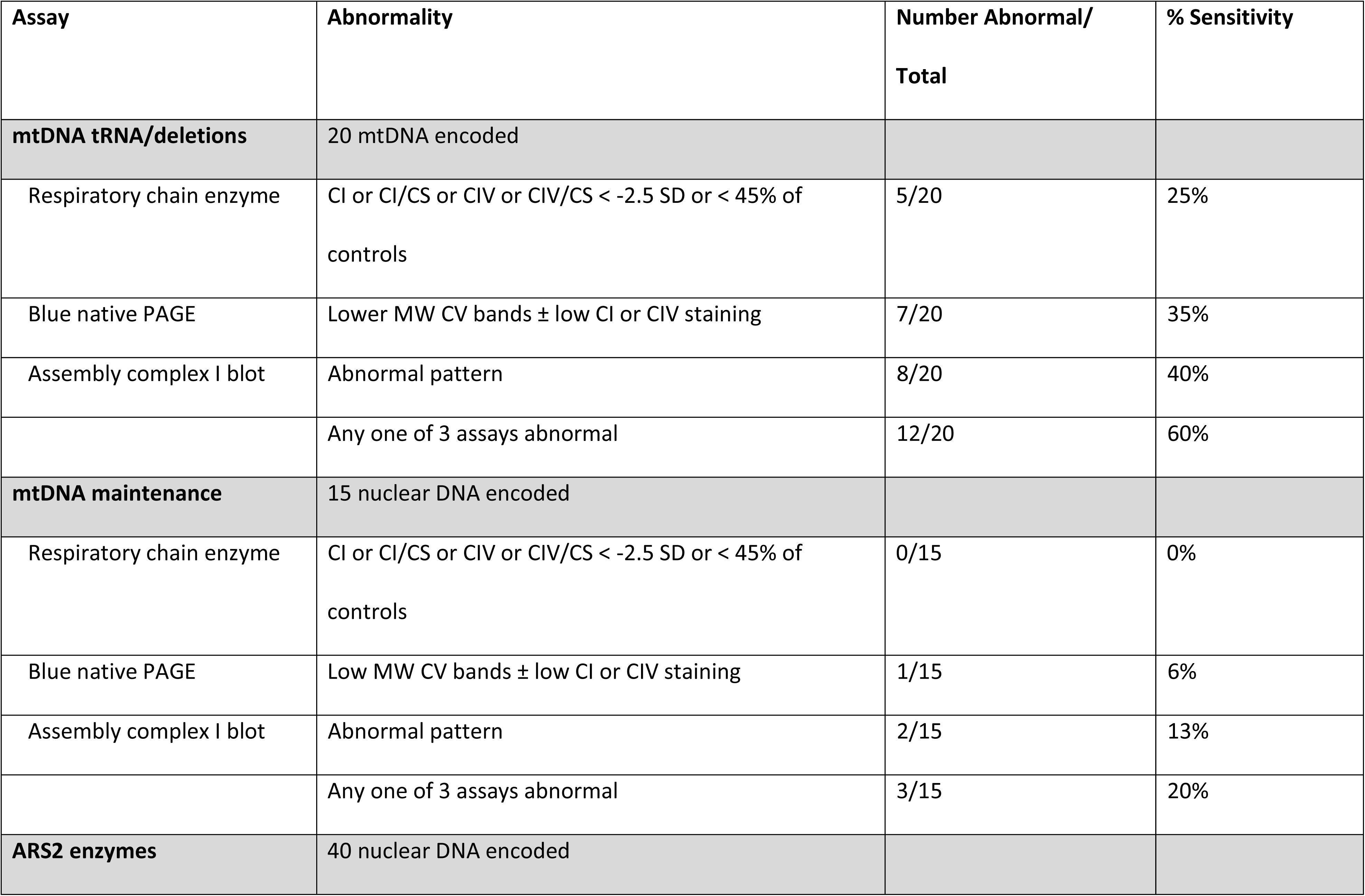
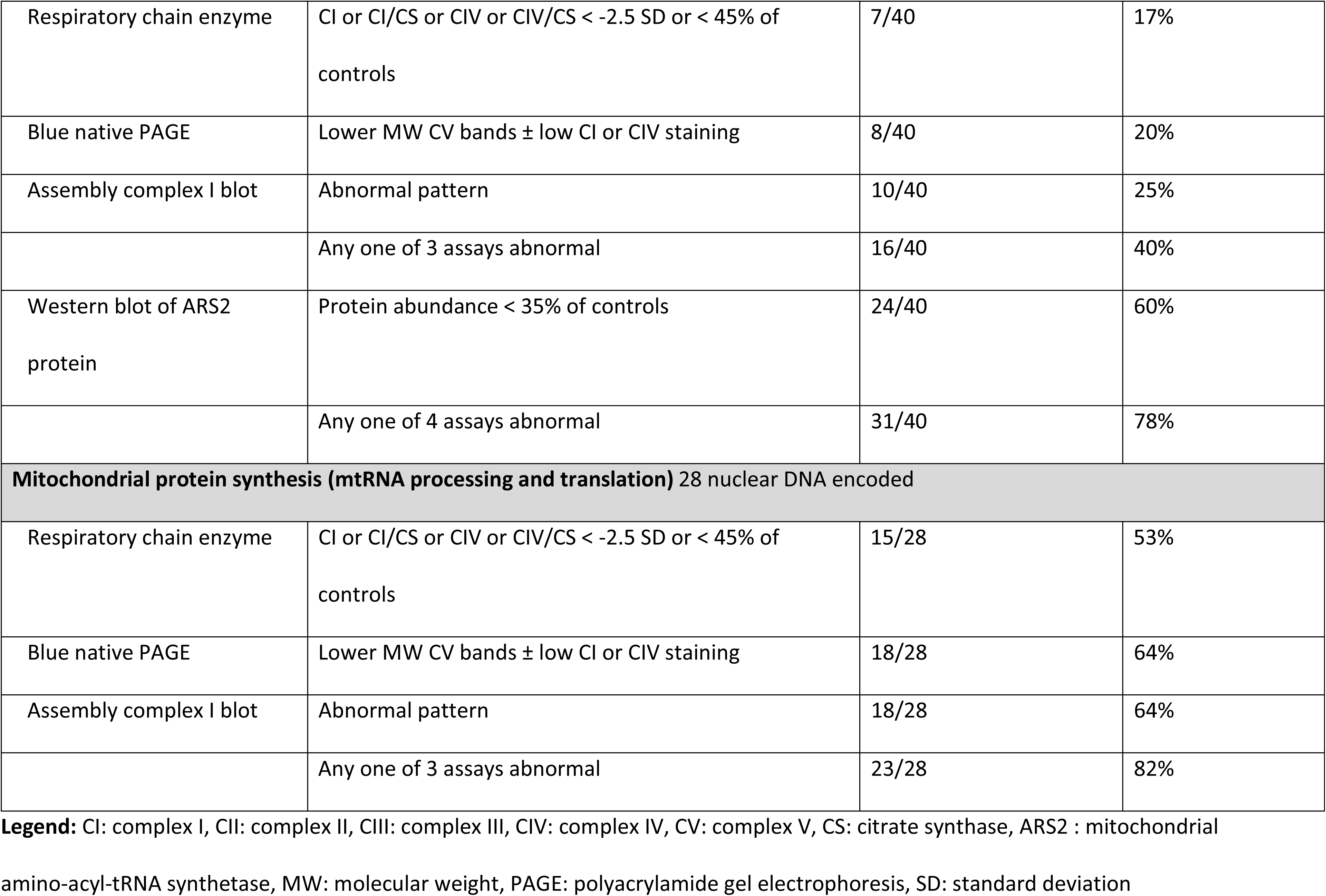
Combined complex deficiencies.

| Assay | Abnormality | Number Abnormal/<br>Total | % Sensitivity |
| --- | --- | --- | --- |
| **mtDNA tRNA/deletions** | 20 mtDNA encoded |  |  |
| Respiratory chain enzyme | CI or CI/CS or CIV or CIV/CS < -2.5 SD or < 45% of controls | 5/20 | 25% |
| Blue native PAGE | Lower MW CV bands ± low CI or CIV staining | 7/20 | 35% |
| Assembly complex I blot | Abnormal pattern | 8/20 | 40% |
|  | Any one of 3 assays abnormal | 12/20 | 60% |
| **mtDNA maintenance** | 15 nuclear DNA encoded |  |  |
| Respiratory chain enzyme | CI or CI/CS or CIV or CIV/CS < -2.5 SD or < 45% of controls | 0/15 | 0% |
| Blue native PAGE | Low MW CV bands ± low CI or CIV staining | 1/15 | 6% |
| Assembly complex I blot | Abnormal pattern | 2/15 | 13% |
|  | Any one of 3 assays abnormal | 3/15 | 20% |
| **ARS2 enzymes** | 40 nuclear DNA encoded |  |  |
| Respiratory chain enzyme | CI or CI/CS or CIV or CIV/CS < -2.5 SD or < 45% of controls | 7/40 | 17% |
| Blue native PAGE | Lower MW CV bands ± low CI or CIV staining | 8/40 | 20% |
| Assembly complex I blot | Abnormal pattern | 10/40 | 25% |
|  | Any one of 3 assays abnormal | 16/40 | 40% |
| Western blot of ARS2 protein | Protein abundance < 35% of controls | 24/40 | 60% |
|  | Any one of 4 assays abnormal | 31/40 | 78% |
| **Mitochondrial protein synthesis (mtRNA processing and translation) 28 nuclear DNA encoded** |  |  |  |
| Respiratory chain enzyme | CI or CI/CS or CIV or CIV/CS < -2.5 SD or < 45% of controls | 15/28 | 53% |
| Blue native PAGE | Lower MW CV bands ± low CI or CIV staining | 18/28 | 64% |
| Assembly complex I blot | Abnormal pattern | 18/28 | 64% |
|  | Any one of 3 assays abnormal | 23/28 | 82% |

**Legend:** CI: complex I, CII: complex II, CIII: complex III, CIV: complex IV, CV: complex V, CS: citrate synthase, ARS2 : mitochondrial amino-acyl-tRNA synthetase, MW: molecular weight, PAGE: polyacrylamide gel electrophoresis, SD: standard deviation

#### mtDNA maintenance

Disorders of mtDNA maintenance demonstrated the lowest overall diagnostic sensitivity. No cases showed an abnormality on RCE, only a single case had an abnormality on BN-PAGE, and only two cases had an abnormal complex I assembly blot. Overall, the combined sensitivity using these assays was only 20% of all cases.

#### Mitochondrial protein synthesis

In contrast, fibroblasts had good sensitivity for disorders of mitochondrial protein synthesis, comprising disorders of mitochondrial transcription, RNA processing and translation and protein import. An abnormality was observed on RCE assay in 53% of cases (nine complex I, four complex IV, and two combined I+IV deficiencies), on BN-PAGE assays was present in 64% of cases, most often as lower MW bands of complex V (15/18), and on complex I assembly assay in 64% of cases. An abnormality on any of the three assays was present in 82% of cases. The five cases with normal assays had pathogenic variants in *MRPL44*, *MRPS16*, *MRPS18C*, *SHMT2*, and *RMND1*. Three of these (*MRPL44, MRPS16*, and *RMND1*) had very low MTCO1 levels, but were normal in *MTPS18C* and *SHMT2*, illustrating that protein levels could be diagnostically useful for this category as well. The diagnostic rate was not significantly different between RNA processing (N=7) and ribosomal translation disorders (N=20) (Chi-square 0.27, p=0.6).

#### ARS2

In the category of the amino-acyl-tRNA synthetase defects, an abnormality on RCE assay was present in only 15% of cases, all but one showed borderline results (between 40 and 45% of average of controls), and on BN-PAGE assay in 20% of cases, and on complex I assembly blot in 25% of cases. An abnormality on any of all three assays was present in 42% of cases. An abnormality on the abundance of the ARS2 protein defined as ≤35% of in-assay controls was present in 60% of cases, giving an abnormality on any of the four assays present in 78% of cases. Of 35 cell lines analyzed, MTCO1 had normal abundance in 23 (66%), strong decrease (<60%) in nine (26%), and borderline decrease (60-67%) in three cases (9%).

Theoretically in these disorders, all complexes that contain mtDNA-encoded subunits should have deficient activity, but this is rarely observed. There were 27 cell lines with deficient RCE activities, including 15 with isolated deficient complex I activity, seven with isolated deficient complex IV activity, and only five with a combined deficiency of complex I and complex IV. For the 22 cell lines with an isolated deficiency on enzymology, the use of both the blue native PAGE and the complex I assembly blot studies provided evidence of another deficient complex in 12 additional cases increasing the evidence of a combined deficiency to 17/22 cell lines.

### Co-factor abnormalities and structural defects **(Table 3)**

The combined complex II-III assay is dependent on endogenous coenzyme Q levels, but this assay was normal in the single coenzyme Q deficient cell line, as previously reported (Rahman et al., 2012). Lipoate is necessary for the activities of PDH and 2-ketoglutarate dehydrogenase. Its synthesis by lipoate synthase (*LIAS*) requires iron-sulfur clusters and SAM as a co-factor. An abnormal PDH enzyme activity and abnormal western blot specific for lipoylated proteins was observed in all four cell lines with disorders of lipoate synthesis, in three of five iron-sulfur clusteropathies, and in the single *SLC25A26* SAM transporter protein deficient cell line. Aconitase enzyme activity, which is dependent on iron-sulfur clusters, was decreased in two of three Fe-S clusteropathies tested (*FDXR* and *NFS1*, the only abnormality in the latter) (Yang et al., 2022). In complex II the iron-sulfur clusters are attached to the SDHB subunit (Bezawork-Geleta et al., 2017). The SDHB abundance on western blot was decreased in *FDXR* and *GLRX5*, and complex II RCE activity was decreased in two cases of *BOLA3* and on BN-PAGE in two cell lines. Complex I contains eight iron-sulfur clusters, and two cell lines had an abnormal complex I assembly blot (*FDXR* and *BOLA3*). This provided an overall sensitivity for the enzyme assays in 10/11 cases (91%) amongst the different enzyme assays, further aided by targeted western blot assays, but was low for BN-PAGE (18%) and complex I assembly blot (18%).

**Table 3:**
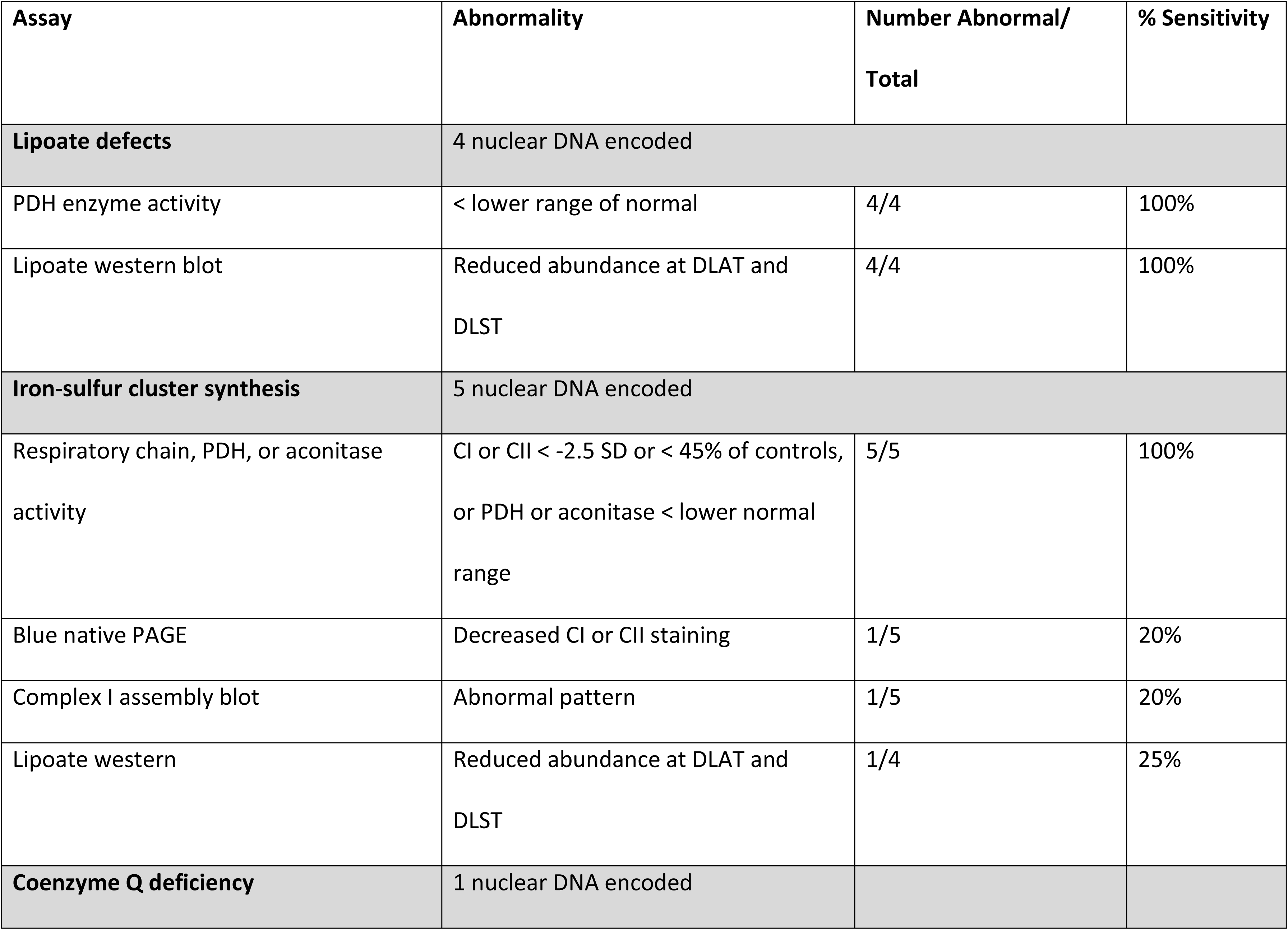
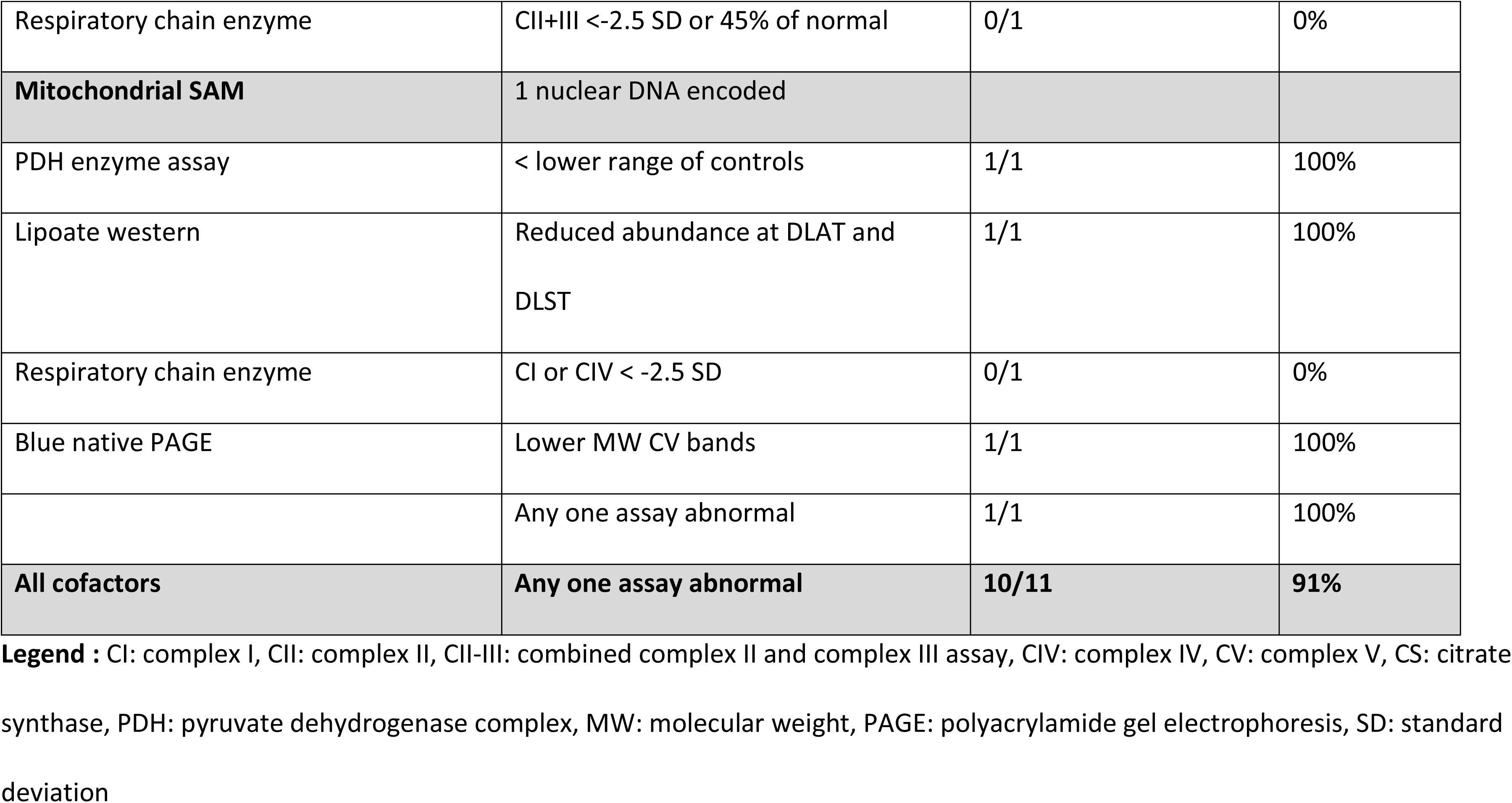
Co-factor deficiencies.

The two cell lines with structural defects (*OPA1*) did not show any functional defects in our system.

### Controls and Differential diagnostic cell lines **(Table 4)**

In control cell lines, no false positives were present on any of the three assays: RCE activities, BN-PAGE or complex I assembly blot. For differential diagnostic cell lines, in the group of the metabolic diseases known secondary abnormalities were identified: reduced PDH activity was noted in one *ECHS1* cell line, reduced complex IV activity was noted in two propionic acidemia cell lines (one of which also had low complex IV activity on BN-PAGE), two pyrophosphatase 2 (*PPA2*) deficient cell lines, and one severe PMM2-CDG cell line. In the myopathies and neuropathy cell lines, no abnormal results were noted. Since frataxin is involved in iron-sulfur cluster biogenesis, it is associated with reduced mitochondrial aconitase activity (Vaubel & Isaya, 2013; Selvanathan & Sankaran, 2022), and this expected finding was indeed present in our cell lines, and was not considered a false positive result. In the genetic syndromes, the cell line with variants in *EIF2AK3* encoding the PERK protein showed low RCE activity of complex IV, on BN-PAGE had low staining for complex I and IV and abnormal low MW bands for complex V, and had an abnormal complex I assembly assay, consistent with a combined defect in mitochondrial complex assembly. PERK function has been associated with mitochondrial membrane biogenesis, supercomplex formation, and is important for mitochondria respiration (Leterme & Michaud, 2023; Sassano et al., 2023; Balsa et al., 2019). No other abnormalities were noted. Thus, the overall false positive rate was 13% for RCE, 4% for BN-PAGE and 2% for complex I assembly assay.

**Table 4:**
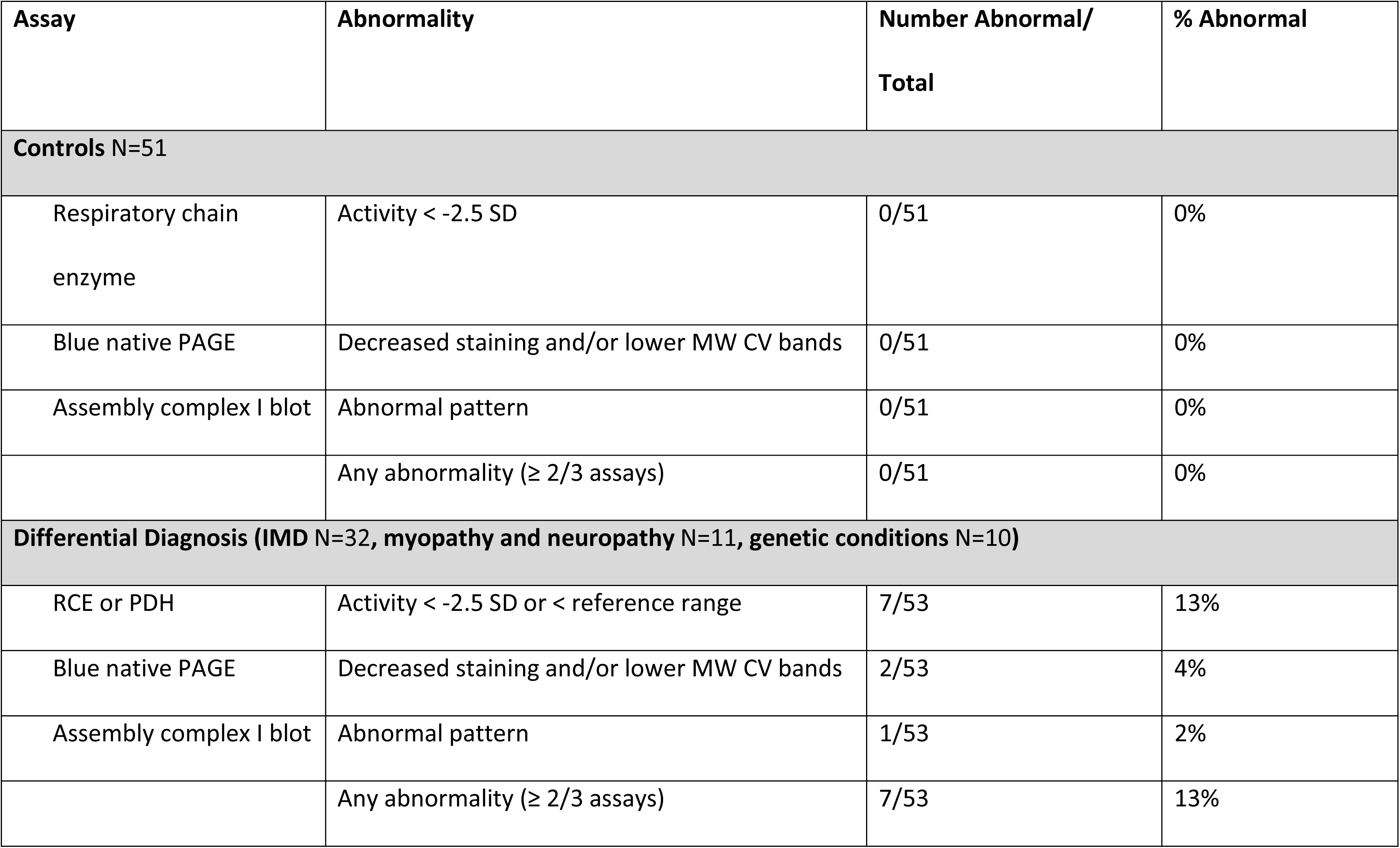
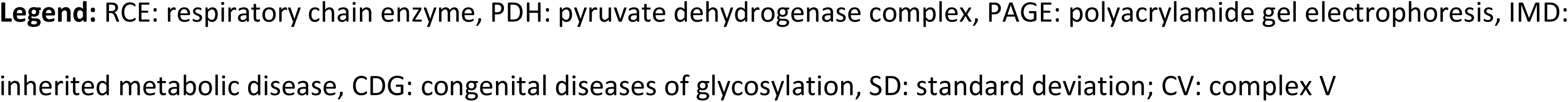
Controls and differential diagnostic conditions.

### Overall analysis (Table 5)

Overall, the sensitivity of diagnostic functional mitochondrial testing collectively for all categories of PMD, as applicable, was 47% for respiratory chain enzyme assays, 40% for BN-PAGE and 49% for the complex I assembly western blot. Combining these three assays had a sensitivity of 68%. The diagnostic sensitivity was further aided by targeted western blot to identify the abundance of specific proteins, which were abnormal in 68% of tested cases, and was most impactful in ARS2 defects. Combining these four functional assays had a sensitivity of 76%, which differed by functional category. The highest sensitivities were for the categories of isolated complex deficiency (85%), mitochondrial protein synthesis disorders (82%) and co-factor defects (91%). In contrast, the lowest sensitivities were observed for the category of mtDNA maintenance disorders (20%), and mtDNA variants (tRNA mutations and deletions) (60%). The sensitivity of diagnosis of ARS2 disorders was markedly increased from 40% to 78% by the addition of western blot-based abundance of ARS2 proteins.

**Table 5:** Combined analysis of clinical utility.

| Category | Total | Enzyme Assays | BN-PAGE | CI assembly | Western | Any Assay Abnormal | % Sensitivity |
| --- | --- | --- | --- | --- | --- | --- | --- |
| **True POSITIVE** | 202 |  |  |  |  |  |  |
| Isolated complex | 88 | 57 | 36 | 35 | 23 | 75 | 85% |
| Mitochondrial protein synthesis | 28 | 15 | 18 | 16 | NA | 23 | 82% |
| Co-factor | 11 | 10 | 2 | 1 | 8 | 10 | 91% |
| mtDNA variants | 20 | 5 | 7 | 8 | NA | 12 | 60% |
| mtDNA maintenance | 15 | 0 | 1 | 2 | NA | 3 | 20% |
| ARS2 | 40 | 7 | 8 | 10 | 24 | 31 | 78% |
| **Total positive/expected** |  | 94/202 | 72/182 | 74/151 | 55/80 | 154/202 |  |
| **Sensitivity** |  | **47%** | **40%** | **49%** | **69%** | **76%** |  |
| **True NEGATIVE** | 55 |  |  |  |  |  |  |
| Differential disease | 53 | 7 | 2 | 1 | NA | 7 |  |
| Controls | 51 | 0 | 0 | 0 | NA | 0 |  |
| **Total false positive** |  | 7 | 2 | 1 |  | 7 |  |
| **ALL COMBINED** |  |  |  |  |  |  |  |
| **Specificity** |  | **93%** | **98%** | **99%** |  | **93%** |  |
| **Positive predictive value** |  | **93%** | **97%** | **99%** |  | **96%** |  |
| **Negative predictive value** |  | **48%** | **53%** | **67%** |  | **67%** |  |

**Legend:** For each category of mitochondrial disorders, and for each control group, the diagnostic performance is provided for the enzyme assays (respiratory chain enzymes, pyruvate dehydrogenase, aconitase), for the blue native polyacrylamide gel electrophoresis (BN-PAGE) with in-gel activity staining, for the complex I assembly assays (CI assembly) and for protein abundance by targeted western blot if indicated. The sensitivities and the specificities are calculated. ARS2 mitochondrial aminoacyl tRNA synthetase.

The specificity was strong. Across all PMDs examined in this study, no false positive results unexpected to the molecular defect were observed. Specificity for RCE activity was slightly decreased (93%) due to the secondary deficiencies observed in certain inborn errors of metabolism, whereas it was high for both the BN-PAGE (98%) and complex I assembly blot (99%), in which the only false positive was seen in *EIF2AK3*. In combination, the specificity of an abnormal result on any test was 93%. Overall, this provides a positive predictive value of 96% for an abnormality on any test (93% for RCE, 97% for BN-PAGE and 99% for complex I assembly assay). The negative predictive value of all assays combined is lower at 67%, reflecting the incomplete sensitivity.

Analysis by sex, excluding X-linked disorders, showed a non-significant higher proportion of positive results in males 78.6% (77 of 98) compared to females 69.9% (65 of 93) (Chi-square = 1.88, p=0.17). Analysis by age showed a lower diagnostic rate in adults 44% (16 of 36) than in children 72% (112 of 155) (Chi-square = 10.2, p=0.0014). This reflects the high prevalence of adults in the poor diagnostic categories of mtDNA maintenance disorders and mtDNA point mutations and deletions, compared to other diagnostic categories.

## Discussion

This study identified the clinical utility in the fibroblast cell lines of a diverse series of primary mitochondrial disorders. It confirmed past retrospective reviews that the sensitivity of fibroblast RCE was only 47% and moreover had a lower specificity of 93% due to secondary mitochondrial dysfunction particularly in select metabolic disorders. However, new assays of BN-PAGE and complex I assembly blot, which had not been systematically studied previously, markedly increased the sensitivity while providing very strong specificity of 98% and 99% respectively. If one considers that this study provides strong confirmatory evidence that *EIF2AK3* constitutes a mitochondrial disorder, as previously suggested (Engelmann et al., 2018), then the specificity becomes near perfect. The complex I assembly blot was particularly effective for nuclear DNA-encoded isolated complex I deficiencies. Adding targeted western blot analysis indicated that protein abundances can help in specific cases. The most striking application is the abundance of ARS2 proteins for ARS2 disorders, which almost doubled the diagnostic rate, and was also beneficial with MTCO1 for mild complex IV deficiency and SDHB for complex II disorders. Overall, these added assays diagnosed an additional 60 patients with a sensitivity increase of 29% without the need for specialized expensive equipment.

Certain categories remain very difficult to diagnose in fibroblasts. The sensitivity is very poor for mtDNA maintenance disorders, as depletion and deletions of mtDNA are usually detected in clinically affected post-mitotic cells. A different approach will be needed for this category. The speed of recovery of mtDNA after its depletion with ethidium bromide or dideoxycytidine could be a candidate approach (Blázquez-Bermejo et al., 2019; Dombi et al., 2024). Sensitivity was also low for mtDNA-encoded variants. However, the impact of this is low as most mtDNA point mutations are now well characterized and rarely require functional assays. One contributing factor may be loss of heteroplasmy in culture; therefore, if pursued, it is important to assay these cell lines in early passages. Our study shows that a combination of all three assays in this category increases sensitivity. Studies at the single cell level such as fluorescent imaging could be an alternative approach to overcome problems associated with the effect of low heteroplasmy (De Paepe et al., 2012). Our study confirms that the ARS2 disorders are difficult to diagnose in fibroblasts as they express few functional defects using traditional techniques. However, we showed that a protein-based approach appeared more promising. Defects of genes involved in the structural maintenance of mitochondria are not recognized on current assays and would benefit from a morphological approach. These poor performing diagnostic categories are highly represented in the adult population, which resulted in overall significantly lower diagnostic sensitivity for adults than for children.

Our study serves as a template to rationally guide clinicians to select the most appropriate assay in a targeted patient-specific way. In general, functional assays are utilized in patients where there is a suspected but not definitive genetic cause: a single variant in a gene known to be recessive, variants of uncertain significance, or candidate genes with known mitochondrial function but not associated yet with a human disease. These genetic candidates guide functional assay selection. For complex I defects, the complex I assembly assay was most effective for nuclear DNA-encoded defects and the RCE was more effective for mtDNA-encoded defects. For mild complex IV defects (e.g. *SCO1* or *SCO2* defects with likely residual function), an added assay to quantify MTCO1 abundance is helpful. For complex V defects a combination of the BN-PAGE and the complex V ATPase hydrolytic assay provided a strong approach for nuclear DNA encoded defects, even for autosomal dominant inheritance mechanisms (Fielder et al., 2025), but was less effective for mtDNA-encoded defects (Friederich et al., 2025). For primary coenzyme Q10 biosynthesis defects, direct measurement of coenzyme Q levels (Yuberro et al., 2014; Rahman et al., 2012) or its synthesis rate (Buján et al., 2014; Tekle et al., 2008) is preferred over reliance on the combined complex II-III assay (Yuberro et al., 2014; Rahman et al., 2012).

This study has two biases. For patients with well-known pathogenic variants, a skin biopsy is rarely taken, thus decreasing the potential sensitivity. Cell lines with variants of uncertain classified as known genetic defects and were thus not included. This increases the sensitivity. A true gold standard does not exist to diagnose disorders such as ARS2 defects, where variants of uncertain significance are frequent. This can theoretically result in an overestimate of the impact of functional tests. We estimate that these two biases likely balanced themselves out in our study.

The diagnostic cutoff was optimized at 45% of average controls based on the data. A Youden index optimization resulted in a cutoff with many false positives in normal control cells (Flkuss et al., 2005). Because the clinical indication for this assay is to support making a definitive diagnosis, this was felt inappropriate for diagnostic purposes, thus we sought to establish a cutoff that maximized specificity over sensitivity. The past arbitrary cutoff of 30% to 40% barely increased specificity but greatly reduced sensitivity (berniet et al., 2002). This is different from results in muscle biopsy, and criteria should be adjusted for this cell type. The diagnostic evaluation established in our laboratory nearly 15 years ago replicated very well in this new set of 51 control cells indicating stability over the years. We did not find an impact of passage number, except perhaps for certain mtDNA defects where heteroplasmy may be lost during prolonged culture. A limitation of our study is that it is retrospective and used samples where the genetics was already known. A prospective study would be useful to determine the predictive value of an abnormal result. Interlaboratory comparisons have highlighted differences in methodology between clinical laboratories performing these mitochondrial functional assays Rodenberg et al., 2013; Chen et al., 2011). In these studies, recognition of complex I and complex III assays was superior in direct isolated assays than in combined linked I-III or II-III assays (chen et al., 2011; chretien et al., 2004), which we also observed. For complex III (Chretien et al., 2004; Birch-Machin et al., 2001), this relates in part to locus of control (Taylor et al., 1994). Provided laboratories use similar isolated complex I and complex III assays, in interlaboratory comparative studies, different laboratories made similar diagnostic discrimination between controls and affected patients despite these small technical differences (Rodenberg et al., 2013; Chen et al., 2011), which were mostly attributed to differences in mitochondrial isolation (Rodenburg et al., 2013). The current study documents the clinical validity of such functional assays.

We conclude that a combination of functional assays including respiratory chain enzyme assay, BN-PAGE and complex I assembly assay provided good sensitivity and excellent specificity for functional mitochondrial studies, sometimes aided with targeted western blot analyses, some of which might in the future be superseded by proteomics studies (Hock et al., 2025). Certain functional groups such as isolated OxPhos complex deficiencies, mitochondrial protein synthesis disorders and co-factor defects have excellent sensitivity in fibroblasts studies. However, in particular mtDNA maintenance disorders are rarely confirmed with current fibroblasts studies. For patients undiagnosed after molecular testing, functional assays in fibroblasts may be a reasonable approach to evaluate for primary mitochondrial disorders.

## Methods

### Fibroblast Cell Lines and culture

#### Cell lines

A case series of fibroblasts was studied between 2014 and 2025 after deidentification under an IRB-approved protocol (COMIRB # 18-1828). They were received from four sources: a local study protocol after obtaining informed consent (COMIRB # 16-0146), after deidentification from the local diagnostic laboratory, provided deidentified from the virtual biorepository of the North American Mitochondrial Disease Consortium (NAMDC) obtained under the NAMDC IRB protocol, or provided from collaborators deidentified under materials transfer agreements. All samples had genetic causes identified by clinical testing, which most often involved next-gen sequencing. Control fibroblasts were obtained deidentified as left-over specimens from routine surgical procedures or from foreskin from circumcision on an IRB-approved protocol (COMIRB # 14-0791). In order to have fair representation of less common categories (e.g. isolated complex II or III deficiencies), a predetermined number of at least 200 PMD patients and at least 100 controls (about half normal and half differential diagnostic category) were prespecified for this study.

#### Tissue culture

Skin fibroblasts were cultured in minimal essential medium-α supplemented with 10% fetal bovine serum, 1X non-essential amino acids and 1X antibiotic/antimycotic solution and cultivated in 5% CO2 at 37 °C, with final expansion using roller bottles culture. Two 850 cm^2^ roller bottles provided sufficient materials for testing of RCE enzyme activities, blue native PAGE with in-gel activity staining, and complex I assembly assays, with additional cells grown when targeted assays were indicated. All fibroblast cell lines were routinely tested for mycoplasma contamination using a PCR-based test supplied by ATCC (Manassas, VA). Fibroblasts were harvested at 80% to 90% confluency and detached using trypsin (0.25%) or scraping. Cells were pelleted by centrifugation at 1000 x g for 10 min, washed with 10 mL 1X DPBS and again pelleted by centrifugation at 1000 x g for 10 min. The cells were washed and pelleted 2 more times and then stored at −80°C until ready to assay. Typically, two 850 cm^2^ roller bottles provided sufficient cells to perform respiratory chain enzyme assays, blue native-polyacrylamide gel electrophoresis (BN-PAGE) with in-gel activity staining, complex I assembly western blot analysis, and various other western blot assays. Additional cells were grown specifically for aconitase enzyme assay, complex V oligomycin-sensitive ATPase activity assay and pyruvate dehydrogenase enzyme assay and growth and harvest details are provided with the assay description.

#### Establishment of normal control fibroblast cell lines

Normal control fibroblast cell lines were derived as described (Vangipuram et al., 2013) with some modifications from foreskins of unaffected neonates or from skin biopsies obtained during routine surgical procedures. The biopsy was sectioned into smaller pieces using a surgical blade and seeded in a T25 flask. The T25 flask containing the sectioned biopsy was incubated 15 to 20 min at 37°C to make sure they were adhered to the flask. Tissue culture media (alpha-MEM supplemented with 20% fetal bovine serum, 1X non-essential amino acids and 1X antibiotic/antimycotic solution) was added and the biopsy pieces were incubated in 5% CO2 at 37°C for 1 to 2 weeks until growth of fibroblasts was evident. Newly derived fibroblasts were then detached using trypsin and scraping and transferred to a new flask and propagated as described previously.

### Enzyme activities

#### Respiratory chain enzyme activities

The activities of the respiratory chain enzyme complexes and citrate synthase were assayed spectrophotometrically at 30°C on a WinCary 300 or 4000 spectrophotometer following published methods with some modifications (Figure 2A) (Rahman et al., 1996; Chatfield et al., 2015; Coughlin et al., 2015; Thorburn et al., 2004). All assays routinely include a normal control fibroblast in the experiment. Fibroblast pellets were resuspended in MegaFB buffer (250 mM sucrose, 2 mM HEPES, 0.1 mM EGTA, pH 7.4) and homogenized two times followed by centrifugation at 600 x g for 10 min at 4°C with retention of the supernatant each time. The supernatants were combined and centrifuged at 14400 x g for 10 min at 4°C to pellet the mitochondria. The pellet was then resuspended in 300 to 400 µL MegaFB buffer and 100 µL was reserved (isotonic fraction). The remaining homogenate was centrifuged again at 14400g for 10 min at 4°C and the pellet was resuspended in 300 µL hypotonic buffer (25 mM KPi, pH 7.2, 5 mM MgCl2). Both the isotonic and hypotonic fractions were subjected to 3 freeze-thaw cycles consisting of 1 min freezing in a dry ice/methanol bath and 3 and 4 min thawing in a room temperature water bath respectively. The isotonic fraction was used for complex III and complex II+III enzyme assays and the hypotonic fraction was used for complexes I, II, IV, and citrate synthase enzyme assays. All assays were incubated for 10 min at 30° C before the initiation of the enzyme reaction. Protein was assayed for both the hypotonic and isotonic fractions using the Bio-Rad protein assay (Bio-Rad, Hercules, CA).

**Figure 2:**
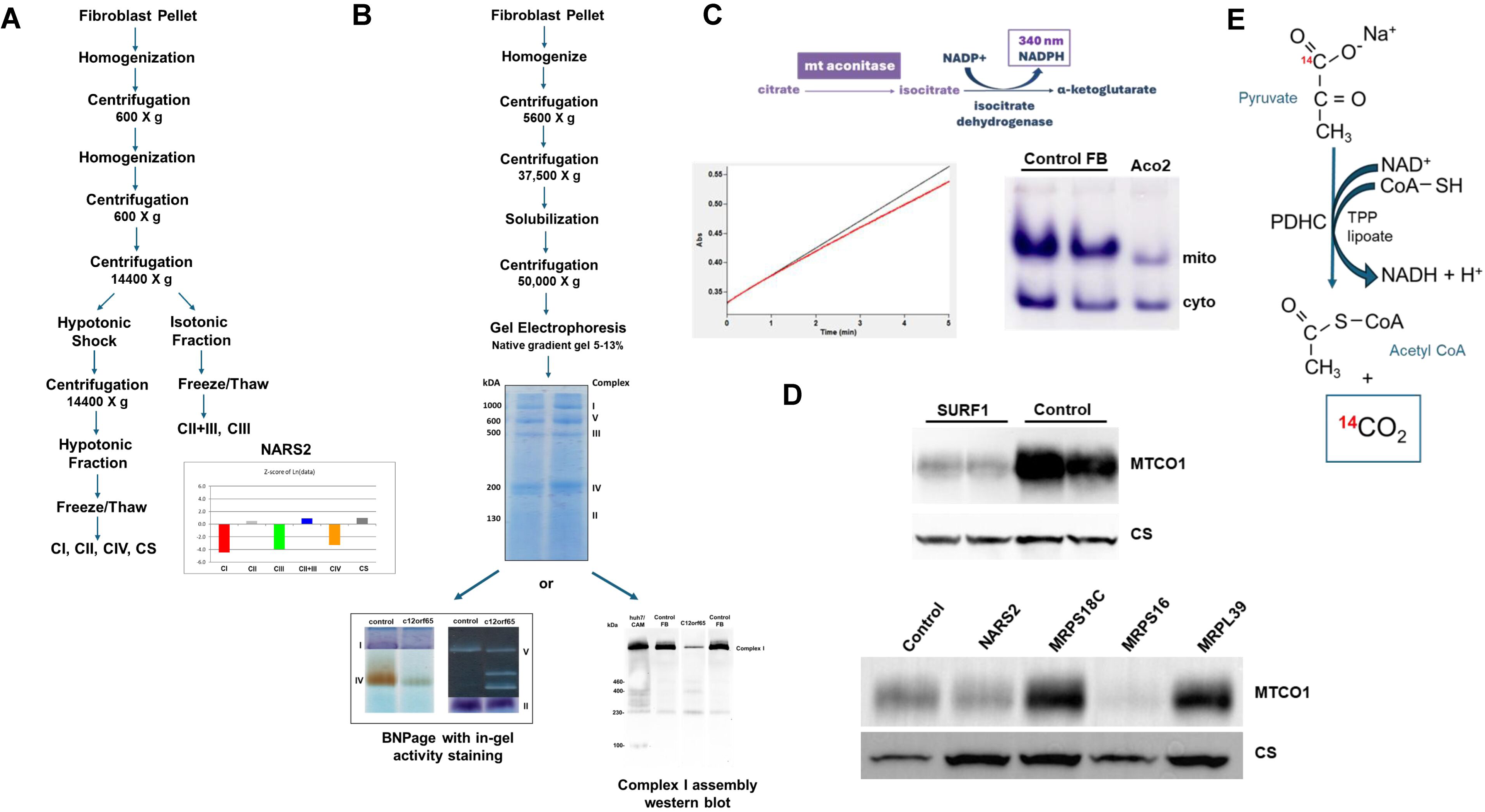
**Schematic presentation of the methods for mitochondrial functional assays**(A). After isolation of mitochondria by differential centrifugation, complex III and II+III are analyzed after a freeze and thaw procedure, whereas the other complexes in addition undergo a hypotonic shock procedure. (B) After isolation of mitochondria by differential centrifugation, the inner mitochondrial membrane proteins are solubilized and separated by blue native gel analysis, with the activities of complexes I, II, IV and V identified by enzymatic colorimetric reaction; left: control, and right: a patient with a combined deficiency. The gel can also be western blotted, probed with an antibody against NDUFS2 to reveal complex I assembly intermediates with normal control (intense band of the holocomplex at 1050 kDa and a faint band at 230 kDa) and a huh 7 cell line treated with chloramphenicol as positive control, and a patient with pathogenic variants in *C12orf65*, which has a decreased holocomplex band, and abnormal intermediates at 400 and 460 kDa. (C). Total aconitase activity is measured by a linked enzymatic spectrophotometric assay (reaction sequence and representative kinetic tracing shown). The mitochondrial and cytosolic aconitase proteins are separated on a non-denaturing gel, and activities revealed by a colorimetric in gel activity staining. A selective decrease in the mitochondrial aconitase activity due to mutations in ACO2 is shown. (D). The abundance of MTCO1 subunit is detected by western blotting of whole cell lysate with citrate synthase as control. Representative decreased abundance in cell lines with pathogenic variants in *SURF1*, *NARS2* and *MRPS16* are shown, whereas the cell lines with variants in *MRSP18C* and *MRPL39* showed normal abundance. (E). The activity of pyruvate dehydrogenase is measured by the release of ^14^C-CO2 from (1-^14^C)-pyruvate.

The activity of complex I was assayed by following the decrease in absorbance at 340 nm due to the oxidation of NADH after incubation with coenzyme Q1. This assay was performed in both the absence and presence of rotenone to account for rotenone insensitive NADH oxidation. For a 500 µL reaction, assay buffer (50 mM KPi, pH 7.4, 1 mM KCN, 0.01 mM antimycin A, 0.1% BSA, 0.05 mM NADH), 0.05 mM coenzyme Q1 and with and without the addition of rotenone (final 2.5 µM) were combined. The reaction was initiated with the addition of 25 µL hypotonic fraction and followed for 3 min at 340 nm. When calculating the activity of complex I, the rotenone-sensitive rate was used which is the difference between the rate determined with the addition of rotenone and the rate without the addition of rotenone. The average percent rotenone sensitivity of the total activity amount in control fibroblast cell lines is 50% (range: 37% to 62%).

The activity of complex II was assayed by following the decrease in absorbance at 280 nm due to the reduction of coenzyme Q1 after incubation with succinate. For a 500 µL reaction, assay buffer (50 mM KPi, pH 7.4, 1 mM KCN, 0.01 mM antimycin A, 2.5 µM rotenone, 10 mM sodium succinate) and 20 µL hypotonic fraction were combined. The reaction was initiated with the addition of coenzyme Q1 (final 0.1 mM) and followed for 3 min at 280 nm.

The activity of complex II + III was assayed by following the increase in absorbance at 550 nm due to the reduction of cytochrome *c*. For a 500 µL reaction, assay buffer (50 mM KPi, pH 7.4, 1 mM KCN, 2.5 µM rotenone, 0.1% BSA, 10 mM sodium succinate) and 10 µL isotonic fraction were combined. The reaction was initiated with the addition of cytochrome *c* (final 50 µM) and followed for 3 min at 550 nm.

The activity of complex III was assayed by following the increase in absorbance at 550 nm due to the reduction of cytochrome *c* after the addition of decylbenzylquinol (Birch-Machin et al., 2001; Rötig et al., 2004). For a 500 µL reaction, assay buffer (50 mM KPi, pH 7.4, 1 mM KCN, 2.5 µM rotenone, 0.1% BSA, 1 mM n-dodecyl-β-D-maltoside) and 7.5 µL isotonic fraction were combined. The reaction was initiated with the addition of decylbenzylquinol (final 50 µM) and cytochrome *c* (final 20 µM) and followed for 5 min at 550 nm. L-ascorbate (final 0.5 mM) was added at the end of the assay and the reaction was followed for an additional 5 min to obtain the final absorbance end point when all cytochrome *c* was reduced.

The activity of complex IV was assayed by following the decrease in absorbance at 550 nm due to the oxidation of cytochrome *c*. For a 500 µL reaction, assay buffer (50 mM KPi, pH 7.4) and 20 µL hypotonic fraction were combined. The reaction was initiated with the addition of reduced cytochrome *c* (final 15 µM) and followed for 5 min at 550 nm. Potassium hexacyanoferrate (final 1 mM) was added at the end of the assay and the reaction was followed for an additional 5 min to obtain the final absorbance end point when all reduced cytochrome *c* was oxidized.

Citrate synthase activity was measured spectrophotometrically by the production of the free sulfhydryl groups (CoASH) using the thiol reagent 5,5-dithio-bis-(2-nitrobenzoic acid) (DTNB) (Matsuoka & Srere, 1973). The chemical coupling of DTNB with the sulfhydryl groups yields a free thionitrobenzoate (TNB) anion. The release of this anion due to reduction of DTNB was measured at 412 nm. For a 500 µL reaction, assay buffer (50 mM KPi, pH 7.4,), acetyl CoA (final 0.1 mM), and 5 µL hypotonic fraction were combined. The reaction was initiated with the addition of oxaloacetic acid (final 0.1 mM) and followed for 3 min at 412 nm.

The enzyme activities for complexes I, II, II+III, and CS were calculated as initial rates using the absorbance difference per minute as calculated by the WinCary program and the specific absorbances coefficient (mM^-1^cm^-1^): 6.81 for NADH; complex I, to account for contribution of coenzyme Q1 reduction to the absorbance at 340 nm; 12.0 for coenzyme Q1 (complex II); 18.7 for reduced cytochrome *c* (complex II+III); 13.6 for TNB product (for citrate synthase). The activities were expressed as nmol/min.mg protein. The enzyme activities for complexes III and IV were calculated as first order rate constants (Fersht 1985). For complex III, the first order rate constant was calculated as detailed, and the end point absorbance was determined by the addition of L-ascorbate (Chatfield et al., 2015; Coughlin et al., 2015). For complex IV, the first order rate constant was calculated using the end point absorbance determined from the complete oxidation of cytochrome *c* with potassium hexacyanoferrate.

The activities of complexes I, II, II-III and citrate synthase were calculated as initial rate, and of complexes III and IV as first order rate constants derived from the first minute of the reaction. They were normalized as a ratio over total cellular protein and as a ratio over the activity of citrate synthase. Normal ranges were derived from 33 control samples (Chatfield et al., 2015; Coughlin et al., 2015Friederich et al., 2017), in which the activities were log normally distributed, allowing expression of the activity as a Z-score on this distribution (DeVore, 2017).

### Normal values

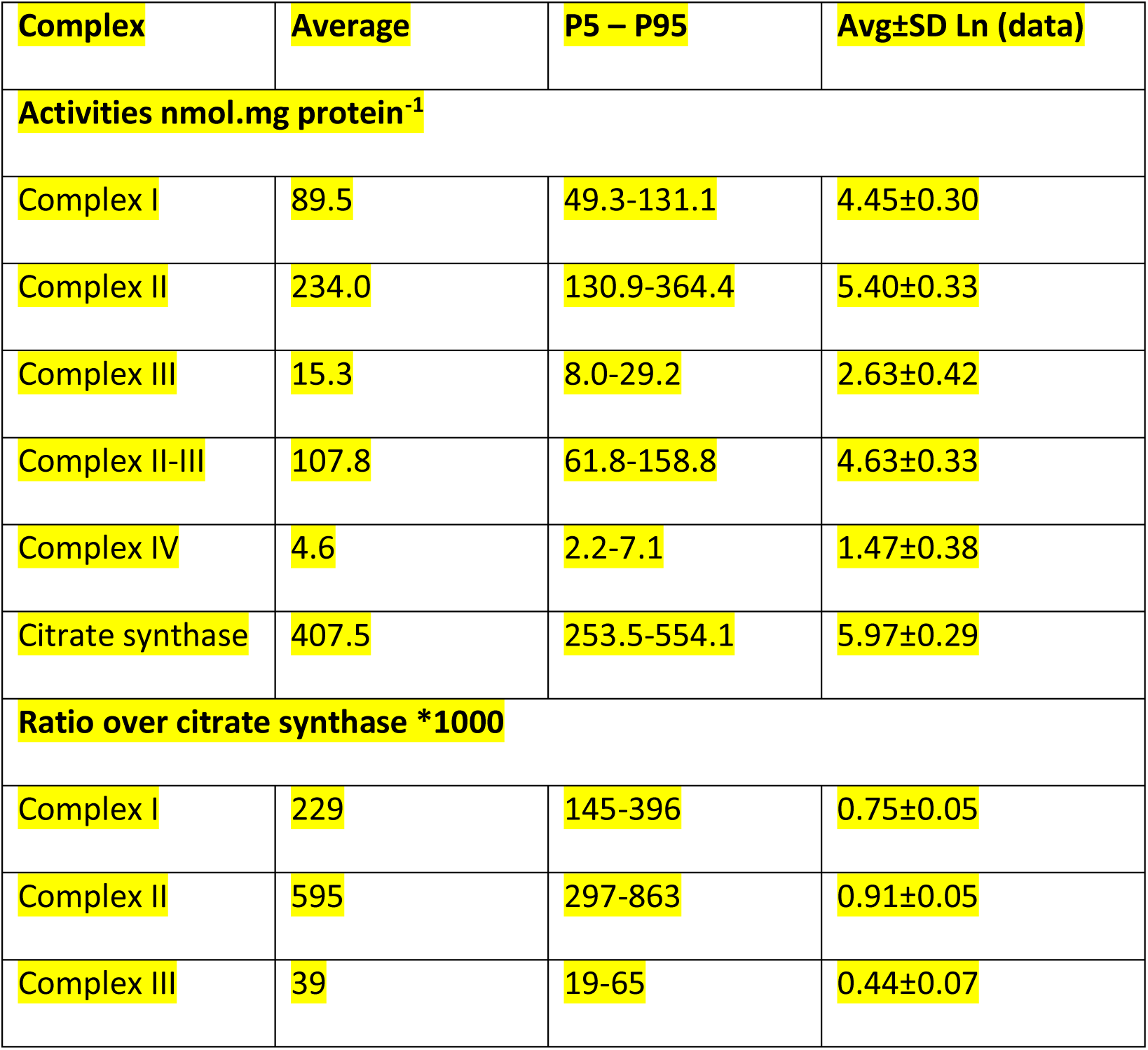
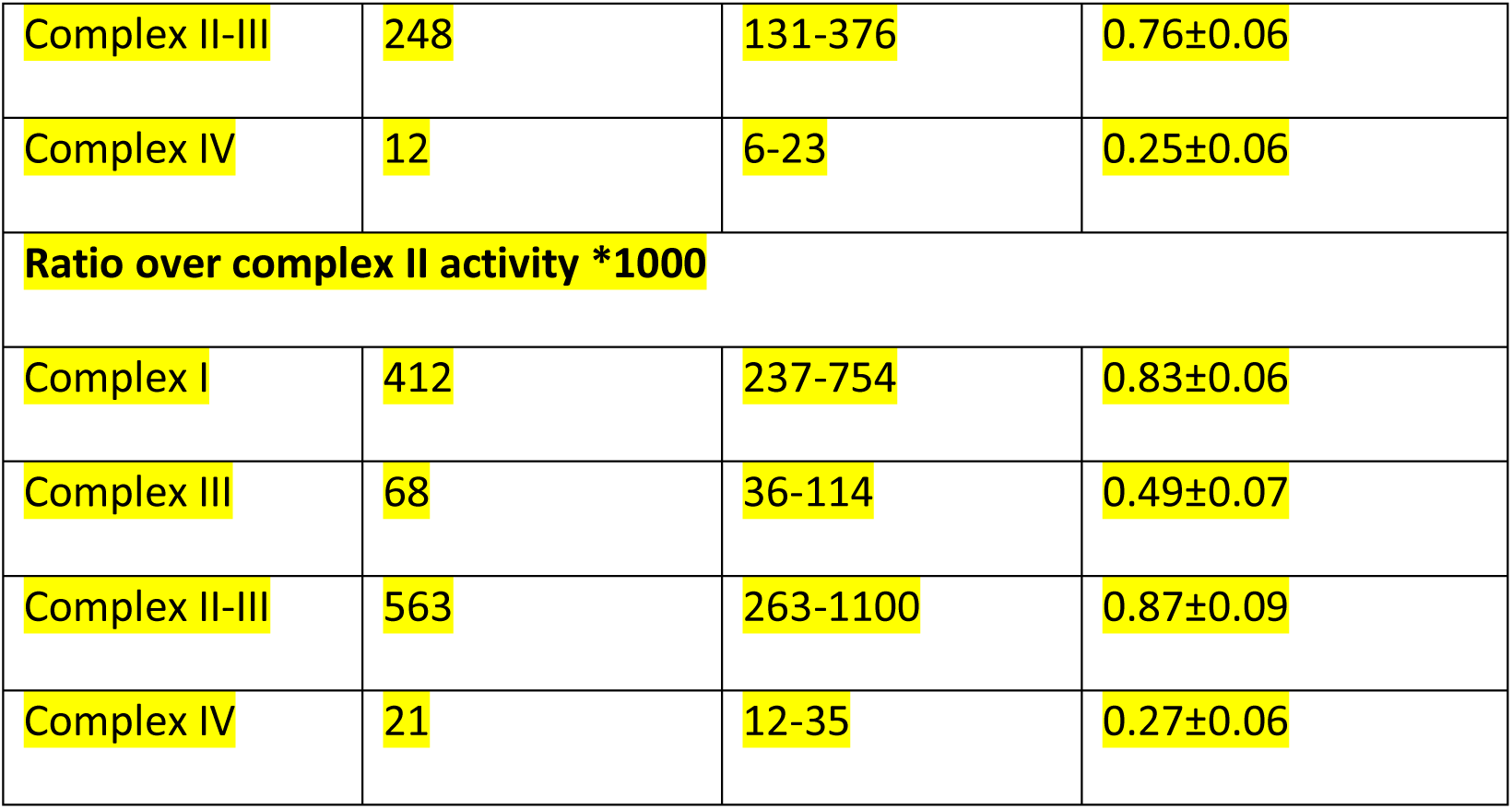

Furthermore, the effect of passage number of fibroblasts on the activities of the respiratory chain enzyme complexes was also evaluated. Cultured skin fibroblasts were harvested at 5, 10, and 15 passages. Respiratory chain enzyme assays were performed, and activities and ratios were calculated as previously described. For the impact of antibiotics and antimycotics, five different cell lines were cultured for three days in parallel with and without antibiotics and antimycotics prior to harvesting, and the respiratory chain enzyme activities measured as above were compared using a paired Student t-test.

### Complex V oligomycin-sensitive ATPase activity assay

Activity of complex V was determined by a kinetic spectrophotometric assay as the oligomycin-sensitive reverse ATP hydrolytic activity using a linked enzymatic assay (Rustin et al., 1994; Friederich et al., 2025; Mayr et al., 2004). In this assay, ATP is converted to ADP and Pi by ATP synthase. ADP generation is coupled via pyruvate kinase and lactate dehydrogenase to the oxidation of NADH to NAD^+^ followed at 340 nm for 5 min at 37°C, after which oligomycin was added and the assay was followed for an additional 5 min to determine oligomycin-insensitive ATPase activity (Friederich et al., 2025).

Fibroblasts were grown in α-MEM supplemented with 10% fetal bovine serum, 1X non-essential amino acids and 1X antibiotic/antimycotic solution. Cells were harvested at 80% confluency from one T150 cm^2^ flask following treatment with trypsin, washed three times with 1X DPBS, and centrifuged at 1000 x g for 10 min after each wash to pellet cells. Cells were assayed immediately after harvesting. The cell pellet was resuspended in MegaFB buffer (250 mM sucrose, 2 mM HEPES, 0.1mM EGTA, pH 7.4), twice homogenized and centrifuged at 1000 x g for 5 min, the supernatant was centrifuged at 14,400 x g for 10 min to pellet the mitochondria and then resuspended in 100 μL MegaFB buffer. Protein was assayed using the Bio-Rad protein assay (Bio-Rad, Hercules, CA). For the ATPase reaction, 50 μL 10X assay buffer (400 mmol/L Tris-HCl, pH 8.0, 50 mmol/L MgCl2, 100 mmol/L KCl, 40 U/mL lactate dehydrogenase, 40 U/mL pyruvate kinase, 2 mmol/L NADH, 20 mmol/L PEP, 5 mmol/L ATP, 1% BSA, 10 μmol/L antimycin A, 3 mM Ap5A, and 30 μmol/L FCCP) and 5 μg protein were combined, and then water pre-warmed at 30°C was added to complete the assay volume to 500 μL. After mixing, it was sonicated for 10 sec using a Branson Sonifier 550 at 20% output and immediately placed in a Cary 4000 UV/VIS spectrophotometer. The oxidation of NADH was followed at 340 nm for 5 min at 37°C, after which 1.2 μL 2.5 mmol/L oligomycin was added and the assay was followed for an additional 5 min to determine the oligomycin-insensitive ATPase activity. The control range for the complex V hydrolysis assay was established from 30 control fibroblast cell lines. All assays routinely include a normal control fibroblast and a known complex V deficient fibroblast in the experiment

### Pyruvate dehydrogenase complex (PDHC) assay

After incubating cells with dichloroacetate, the enzyme activity of pyruvate dehydrogenase complex (PDH) was determined by the release of ^14^C-CO2 from 1-^14^C-pyruvate as described (Figure 2E) (Rahman et al., 1996; Hyland et al., 1983; Wicking et al., 1986; Ferdinandusse et al., 2015). In this PDHC assay the fibroblast homogenate was incubated with [1-^14^C]-Na-pyruvate and the necessary cofactors and evolved radioactive CO2 that is released from the reaction fluid was quantitated by beta-scintillation counter. Fibroblasts were grown in alpha-MEM supplemented with 10% fetal bovine serum, 1X non-essential amino acids, and 1X antibiotic/antimycotic at 37°C with 5% CO2. Cells were harvested at 80 to 90% confluency from two T75 cm^2^ flasks for duplicate assays following treatment with trypsin, washed three times with 1X DPBS, and centrifuged at 1000 x g for 10 min after each wash to pellet cells. The cell pellet was resuspended in 500 µL of 0.5 mM DCA in 1X DPBS followed by incubation at 37°C for 15 min in a shaking water bath.

Treatment with DCA inactivates the kinase which inactivates pyruvate dehydrogenase by phosphorylation. The cells were centrifuged at 1000 x g for 10 min and the pellet was snap frozen on dry ice and stored at −80°C until assayed.

The cell pellet was resuspended in 300 µL PDH homogenization buffer (2.95 mL incomplete buffer: 10 mM Tris-HCl pH 7.4, 50 mM KCl, 4 mM MgCl2, 30 mM nicotinamide, 1 mM Na2SO4, 1 mM EDTA; 50 µL 0.1 M DTT; and 2 mL glycerol) and subjected to 3 freeze-thaw cycles consisting of 1 min freezing in a dry ice/methanol bath and 3 min thawing in a room temperature bath followed by vortexing each time.

For this assay, two normal control cell lines, one PDH-deficient cell line, and a blank sample prepared from boiling cellular extract for 5 min were assayed in addition to patient samples. Reaction vessels with side arms were assembled each containing a center well with 200 µL 10% KOH. Fibroblast cellular extracts were incubated with [1-^14^C]-Na-pyruvate (2.5 µCi/mL) and assay buffer containing the necessary cofactors (40 mM 0.5 M KPi, pH 7.4, 1.3 mm EDTA, 2.5 mM MgCl2, 8 mg/mL BSA, 1.6 mM NAD^+^, 0.9 mM TPP, 0.5 mM DCA, 1% v/v rat serum, 1 mM DTT, 0.15 mM Coenzyme A, 1.0 mM Na2SO3) in a total volume of 500 µL at 37°C for 90 min in a shaking water bath.

The reaction was quenched by the addition of 500 µl 25% TCA and incubated at 37°C for an additional 60 min in a shaking water bath to ensure all evolved CO2 was trapped in the KOH well. The evolved radioactive CO2 was then released by acidification with acetic acid and quantified using a beta-scintillation counter. Protein was assayed using the Bio-Rad protein assay (Bio-Rad, Hercules, CA) and the activity of the PDH complex was calculated in nmol/min/mg protein using the specific activity (dpm/nmol) of [1-^14^C]-Na-pyruvate determined every time an assay is performed. The PDHC enzyme activity control range (0.87-3.13 nmol/min/mg protein) was established from 20 normal fibroblast cell lines. All assays routinely include a normal control fibroblast and a known deficient fibroblast in the experiment.

### Aconitase assay

The aconitase enzyme assay used is a combination of a spectrophotometric kinetic enzyme assay that measures total cellular aconitase activity (cytosolic and mitochondrial enzyme activity) and a non-denaturing PAGE in-gel activity staining assay that allows the separation of the cytosolic and mitochondrial aconitase enzyme activities (Figure 2C) (Fansler & Lowenstein, 1969; Condò et al., 2010; Vögtle et al., 2018; Tong et al., 2006; Baker at al., 2014). Fibroblasts were grown in alpha-MEM supplemented with 10% fetal bovine serum, 1X non-essential amino acids, and 1X antibiotic/antimycotic at 37°C with 5% CO2. Cells were harvested from two to four T150 flasks at 90% confluency, washed three times with 1X DPBS, centrifuged at 1000 x g for 10 min after each wash to pellet cells, and frozen at −80°C until ready to assay.

For the spectrophotometric total enzymatic assay (Condò et al., 2010), cell pellets were resuspended in 2X the volume of cell pellet in triton/citrate lysis buffer (40 mM KCl, 25 mM Tris-Cl, pH 7.5, 1% Triton X-100, 1X AEBSF protease inhibitor, 1.0 mM DTT, 2.0 mM Na citrate, 0.6 mM MnCl2), incubated on ice for 10 min and then centrifuged at 600 x g for 5 min at 4°C retaining the supernatant for the enzyme assay. Protein was assayed using the Bio-Rad protein assay (Bio-Rad, Hercules, CA). For the aconitase spectrophotometric enzyme assay, 50 µL 10X assay buffer (1 M Tris-HCl, pH 7.5, 50 mM NaCitrate, 25 mM MgCl2), 25 µL cellular extract, 2 units/mL isocitrate dehydrogenase, and H2O to bring the assay volume to 500 µL were combined and incubated at 30°C for 5 min. The reaction was then initiated by the addition of 4 µL NADP^+^ (200 mM, final concentration 1.6 mM) and followed for 5 min at 340 nm on a Cary 300 UV/Vis spectrophotometer. The activity of total aconitase enzyme was calculated using the molar extinction coefficient for NADPH (6.22 mM^-1^cm^-1^) (Fansler & Lowenstein 1969). Normal values are 10.14±0.81, range 8.92-13.74 nm.min^-1^.mg protein^-1^.

For the in-gel activity staining assay, 25 µg protein of the triton/citrate lysate was electrophoresed on an 8% polyacrylamide tris borate / sodium citrate gel which separated the mitochondrial from the cytosolic aconitase (Vögtle et al., 2018; Tong et al., 2006; Baker at al., 2014). To 2X sample buffer (1.0 M Tris-Cl pH 8.0, 40% glycerol, 0.1% bromophenol blue) was added to 25 µg cellular extract and separated on an 8% polyacrylamide tris borate NaCitrate gel, 1X tris glycine running buffer (10X running buffer: 250 mM tris base, 1.92 M glycine, pH 8.0) with 3.6 mM NaCitrate added just before use at 4°C for 2 to 2.5 hrs at 160V, < 30 mA. The gel was then incubated in 10 mL activity stain (100 mM Tris-HCl, pH 8.0, 1 mM NADP^+^, 2.5 mM cis-aconitate, 5 mM MgCl2, 1.2 mM MTT, 1 mg PMS, 5 units/mL isocitrate dehydrogenase) for 1 hr at 37°C in the dark and then rinsed 2 to 3 times with H2O, which produces colored bands corresponding to the mitochondrial and cytosolic aconitase activities. The bands corresponding to the mitochondrial aconitase and the cytoplasmic aconitase were then quantified by densitometry, using Bio-Rad Quality One software, and the proportional band intensities were then applied to the enzymatically determined overall activity to calculate the mitochondrial and the cytosolic aconitase activities (Vögtle et al., 2018). The normal control range for this assay was established from eight normal fibroblast cell lines.

### Blue native PAGE with in-gel activity staining

Mitochondrial membrane fractions were isolated by differential centrifugation, solubilized, and the proteins were separated by non-denaturing blue native polyacrylamide gel electrophoresis (BN-PAGE) (Figure 2B) (Chatfield et al., 2015; Coughlin et al., 2015; Van Coster et al., 2001; Smet et al., 2005; Smet et al., 2009). The gel was then cut corresponding to the locations of each complex, and the enzyme activity of each complex was determined by an in-gel activity staining. The fibroblast pellet was resuspended in 2 mL Zheng buffer (210 mM mannitol, 70 mM sucrose, 5 mM HEPES, 1 mM EGTA, pH 7.2) and transferred to a homogenizer. The sample was disrupted with 30 strokes using a Teflon pestle until evenly dispersed and centrifuged at 5600 x g for 1 min at 4°C. The supernatant was retained, and the resulting pellet was resuspended in 2 mL Zheng buffer, homogenized, and centrifuged as described previously. The supernatants were combined and centrifuged at 37,500 x g for 3 min to pellet the mitochondria. The mitochondrial pellet was resuspended in 1 mL Zheng buffer, homogenized as described previously, and centrifuged at 37,500 x g for 3 min. The pellet was washed with 500 µl Zheng buffer and centrifuged for 1 min at 5600 x g. The supernatant was retained and then centrifuged again at 10,000 x g for 10 min. The final pellet containing the mitochondria was stored at −80°C. The isolated mitochondria pellet was thawed on ice and then solubilized in 96 µL 750 mM aminocaproic acid, 50 mM BisTris, 20 μM PMSF, and 12 µL 10% n-dodecyl-β-D-maltoside, vortexed, and centrifuged at 50,000 x g for 30 min at 4°C to pellet mitochondrial particulate material. The clear supernatant containing the oxidative enzyme complexes was retained and stored at −80°C until use.

Protein was assayed using the Bio-Rad protein assay (Bio-Rad, Hercules, CA) and 30 µg protein in 1X sample loading buffer (20X sample loading buffer: 5% Serva Blue G, 750 mM aminocaproic acid) was loaded in duplicate lanes on a 5 to 13% polyacrylamide non-denaturing gradient gel made with 1X gel buffer (3X gel buffer: 1.5 M aminocaproic acid, 150 mM Bis-Tris) and a 40% acrylamide/bisacrylamide mixture (41.25% T, 3% C). The gel was run at 4°C at 75V using 1X anodal buffer (5X anodal buffer: 250 mM Bis-Tris, pH 7.0) in the lower chamber and colored cathode buffer (50 mM Tricine, 15 mM Bis-Tris, 0.2% Serva G, pH 7.0) in the upper chamber of the gel electrophoresis apparatus. After 1 hr, the colored cathode buffer was replaced with cathode buffer (50 mM Tricine, 15 mM Bis-Tris, pH 7.0) and electrophoresis was continued for an additional 1.5 to 2 hrs at 200V until the Serva G tracking dye ran off the gel.

The gel was then cut into four sections corresponding to the positions of migration of complex I, complex II, complex IV, and complex V. The sectioned gels were placed in the appropriate staining solution and incubated at 37°C: complex I (2 mM Tris-HCl, pH 7.4, 0.1 mg/ml NADH, 2.5 mg/ml nitro blue tetrazolium (NBT)) for 1 to 2 hrs; complex II (4.5 mM EDTA, 10 mM KCN, 0.2 mM phenazine methosulfate (PMS), 84 mM Na succinate, and 10 mM NBT in 1.5 mM phosphate buffer, pH 7.4) for 2 to 3 hrs; complex IV (5 mg 3,3’-diaminobenzidine (DAB) in 9 ml 0.05 M phosphate buffer, pH 7.4, 1 ml catalase (20 µg/ml), 10 mg cytochrome *c*, and 750 mg sucrose) overnight; and complex V (35 mM Tris-HCl, 270 mM glycine, 14 mM MgSO4, 0.2% Pb(NO3)2, and 8 mM ATP, pH 7.8) for 4 to 5 hrs. After incubation, the gels were fixed for 15 min in 50% methanol and 10% acetic acid except for the gel assayed for complex V which was rinsed with water and all gels were scanned to obtain images.

This assay is a qualitative assay therefore a normal fibroblast cell line is assayed in tandem with the patient cell line for comparison. In this assay, an abnormal result was visualized as reduced or absent activity staining, or for complex V as bands of lower MW which are observed in disorders affecting complex V, or in disorders of mtDNA maintenance, transcription or translation (Smet et al., 2009).

### Complex I assembly blot

To trace the assembly of complex I, solubilized inner mitochondrial membrane fraction prepared as for the BN-PAGE was separated on a non-denaturing polyacrylamide gel, followed by western blot transfer to a PVDF membrane, decorated using an antibody against NDUFS2, a subunit of the earliest assembly stage of complex I, and detected with a secondary antibody using enhanced chemiluminescence (Figure 2B) (Friederich et al., 2017). In a normal control, this assay detects the holocomplex at 1,050 kDa as a thick band, and in some controls, it also detects the presence of a 230 kDa subcomplex. The intensity of the 230 kDa band obtained by densitometry was <18% of the intensity of the holocomplex in all normal fibroblasts analyzed. An abnormal result is a decrease in the intensity of the holocomplex, an increase in the intensity of the 230 kDa intermediate, or visual presence of intermediate assembly bands at 100, 400, 460, or 950 kDa. Negative and positive controls were consistently run in the same assay.

Solubilized mitochondrial inner membrane preps were isolated as detailed for the BN-PAGE with in-gel activity staining assay. 20 µg protein was loaded on a 5 to 13% polyacrylamide non-denaturing gradient gel and electrophoresed as described previously. Following electrophoresis, the gel was transferred to an Immobilon-P membrane after equilibration in T-buffer (48 mM Tris, 38 mM glycine, 1.3 mM SDS, 20% methanol). After transfer, the membrane was blocked with 1X PBS, pH 7.4 containing 0.2% Tween-20 and 5% non-fat milk for 2 hrs at room temperature and rinsed thoroughly with water to remove residual milk. The membrane was incubated overnight at 4°C with an antibody specific for NDUFS2 diluted 1:2000 in 1X PBS, pH 7.4 with 3% BSA and 0.02% thimerosal. The membrane was washed 4 times with 1X PBS, pH 7.4 with 0.2% Tween-20 for 10 to 15 minutes each time with shaking and then incubated with the appropriate HRP-conjugated secondary antibody for 1 hr at room temperature. The membrane was finally washed 2 times 1X PBS, pH 7.4 with 0.2% Tween-20 followed by 2 times with 1X PBS, pH 7.4 and visualized by enhanced chemiluminescence. For this assay, a control fibroblast cell line was assayed as a negative control while a HepG2 or Huh7 cell line grown in DMEM with 40 μg/ml chloramphenicol for 48 hours was used for a positive control.

### Protein abundance by Western blots

Protein abundance by western blot was determined of SDHA and SDHB for isolated complex II deficient cases, of MTCO1 (a.k.a. COX1) for complex IV deficient cases and mitochondrial translation deficient cases (Figure 2D) (Kripps et al., 2020), of ATP5F1A and ATP5PO for isolated complex V deficient cases Ganapathi et al., 2022), and of each aminoacyl tRNA synthetase ARS2 protein in all ARS2 deficient cases. For lipoate deficiency disorder a western blot specific for lipoylated proteins DLAT and DLST was used Baker et al., 2014).

Western blot analysis was performed on mitochondria isolated by differential centrifugation from cultured skin fibroblasts or on remaining homogenates from various enzyme activity assays. Each experiment included three controls. 10 to 20 µg protein in 1X sample loading buffer (2X Bio-Rad Laemmli Sample Buffer with 2-mercaptoethanol) was loaded on varying percentage of polyacrylamide SDS gels dictated by the molecular weight of the protein being probed and electrophoresed at 100 to 150V for 1 to 2 hrs using 1X running buffer (10X 25 mM Tris, 192 mM glycine, 0.1% SDS). Following electrophoresis, the gel was transferred to an Immobilon-P membrane after equilibration in T-buffer (48 mM Tris, 38 mM glycine, 1.3 mM SDS, 20% methanol). After transfer, the membrane was blocked with 1X PBS, pH 7.4 containing 0.2% Tween-20 and 5% non-fat milk for 2 hrs at room temperature and rinsed thoroughly with water to remove residual milk. The membrane was incubated overnight at 4°C with the appropriate antibody diluted in 1X PBS, pH 7.4 with 3% BSA and 0.02% thimerosal. The membrane was washed 4 times with 1X PBS, pH 7.4 with 0.2% Tween-20 for 10 to 15 minutes each time with shaking and then incubated with the appropriate HRP-conjugated secondary antibody for 1 hr at room temperature. The membrane was finally washed 2 times using 1X PBS, pH 7.4 with 0.2% Tween-20 followed by 2 times with 1X PBS, pH 7.4 and visualized by enhanced chemiluminescence. Protein bands of interest were quantitated using ImageJ software and normalized to either citrate synthase or adenine nucleotide translocase (ANT).

#### Lipoic Acid

Lipoylation of the E2 components of pyruvate dehydrogenase DLAT and α-ketoglutarate dehydrogenase DLST was evaluated by western blotting following electrophoresis on a 10% polyacrylamide SDS gel of PDH enzyme assay homogenates. The samples were probed using an anti-lipoid acid antibody (Baker et al., 2014).

#### mtARS2

The ARS2 proteins were analyzed by western blotting following polyacrylamide SDS gel of isolated mitochondrial membrane fractions, or by using a solubilization of the whole fibroblast pellet in RIPA buffer (Thermo Scientific # 89901). Using a mitochondrial membrane fraction for these assays reduced the appearance of non-specific bands.

#### MTCO1

This protein is very sensitive to mitochondrial translational defects and due to the hydrophobic nature of this protein, samples are not boiled prior to loading on the SDS gel. Enzyme assay homogenates were evaluated by western blotting following electrophoresis on a 10% polyacrylamide SDS gel.

#### ATP5PO, ATP5F1A, ATP5F1B

The integrity of complex V was evaluated using a combination of antibodies specific for proteins of this complex: ATP5PO, the linker between the F1 catalytic core and the peripheral stalk; ATPF1A, the α subunit of the F1 catalytic core; and ATPF1B, the β subunit of the F1 catalytic core Ganapathi et al., 2022). Enzyme assay homogenates were evaluated by western blotting following electrophoresis on a 10% polyacrylamide SDS gel.

#### SDHA and SDHB

Patient cell lines with complex II defects were evaluated using antibodies specific for two subunits of this complex: SDHA and SDHB. Enzyme assay homogenates were evaluated by western blotting following electrophoresis on a 10% polyacrylamide SDS gel.

A list of reagents, a list of antibodies and dilutions, and a list of additional method abbreviations is provided in Appendix Table 1.

### Data analysis

For all samples, age, sex, and genotype were recorded. For mtDNA variants, the heteroplasmy rate provided in blood was recorded, and the heteroplasmy rate in the fibroblasts under study was determined at the Children’s Hospital Colorado Precision Medicine clinical laboratory from their whole genome sequencing studies. Laboratory assays and scoring of results was done blinded to the genetic cause.

For descriptive analysis, data were discretized to normal or abnormal test result and compared to the expected result for the molecular disease category. For instance, an abnormal complex I assembly assay would be scored as a false positive result for a patient with a primary isolated complex IV deficiency. For each patient category, the expected test result was thus categorized, which increased the sample number in the control category for specific assays. For optimization of the diagnostic cutoff, the number of abnormal test results were analyzed across a range of cutoffs for both the percent of the mean of normal controls and for the Z-score. Sensitivity, specificity, and positive and negative predictive values were calculated for each test. The analysis of all primary mitochondrial disorders for differences in sensitivity by age (children versus adults) or by sex, excluding X-linked disorders (*PDHA1*, *AIFM1*), was evaluated by Chi-square test.

## Data Availability

The data supporting the findings of this study are available within Tables EV1, EV2, and EV3. Additional materials are deposited in Zenodo available at 10.5281/zenodo.20615940. The data can be reviewed at https://zenodo.org/records/20615940?preview=1&token=eyJhbGciOiJIUzUxMiJ9.eyJpZCI6ImRjNTdkNDE3LTNiNGEtNGI3YS1hYWI0LWU0ZjhiZjdiNzZjYiIsImRhdGEiOnt9LCJyYW5kb20iOiI5ZmVjOGE3NjhjNTI0NDA0NzNiZTQ2ZGFhMWRjNTgyOSJ9.c3qdC7fXuOPUFhOwZuO14Yq5ncFv9qjm9Nh-Q1caNpnF94dFQSwPejHj9U7zk0Cnj20CgS3ixuwt07hcjIxv4A.

## Expanded view

Expanded view contains 4 tables.

## Appendix Tables

There are 3 appendix tables.

Appendix document: Strobe Statement

## Contributions

Study concept and design: JVH, MWF; Laboratory studies: MWF, RAVH, JCL, KMK, TED; Statistical analysis: JVH, LS; Sample and data contributions: JVH, RDG, MH, AAL, RPS, AG, MT, RHH, UL-K, JVH, BHC, RNAVC, JEGS, AVV, DC, PV, JAM, SBW, LAW, WAG, AK, KW, MGB, EM, GCG, WES, SHE, JA, MS, JAR, CS-A, JTP, RG, GME, ES, FS, KAK, EEG, JHY, ANL. First draft writing: JVH, MWF; Critical rewriting: JVH, MWF, LS, RDG, MH; Final responsibility, guarantor, and communicating author: JVH. All authors have seen, reviewed, and approve of the final manuscript.

## Acknowledgements

Funding: This study was supported by a grant from the National Institutes of Health, NIH U54NS078059 for the North American Mitochondrial Disease Consortium (NAMDC) to J.L.K.V.H. NAMDC is part of Rare Diseases Clinical Research Network (RDCRN), an initiative of the Office of Rare Diseases Research (ORDR), NCATS. Contents are the authors’ sole responsibility and do not necessarily represent official NIH views. It also received philanthropic support from the Children’s Hospital Colorado Summits for Samantha, and the University of Colorado Foundation. All funding sources had no role in the design or execution of the study, the interpretation of data, or the writing of the study.

This research was supported in part by the Intramural Research Program of the National Institutes of Health (NIH). The contributions of the NIH authors (LAW, WAG) are considered Works of the United States Government. The findings and conclusions presented in this paper are those of the authors and do not necessarily reflect the views of the NIH or the U.S. Department of Health and Human Services. One of the authors of this publication (SBW) is a member of the European Reference Network for Rare Hereditary Metabolic Disorders (MetabERN) – Project ID No 739543.”

## Abbreviations

ANOVA: analysis of variance
ARS2: mitochondrial amino acyl-tRNA synthetase
BN-PAGE: blue native- polyacrylamide gel electrophoresis
mtDNA: mitochondrial DNA
NADH: nicotine adenine dinucleotide (reduced)
NAD^+^: nicotine adenine dinucleotide (oxidized)
NAMDC: North American Mitochondrial Diseases Consortium
nDNA: nuclear DNA
next-gen: next generation sequencing
OxPhos: oxidative phosphorylation
PAGE: polyacrylamide gel electrophoresis
PDH: pyruvate dehydrogenase complex
PMD: primary mitochondrial disease
RCE: respiratory chain enzyme
SAM: S-adenosylmethionine
VUS: variant of uncertain significance

